## Supplementary Figures 1-18 and Supplementary Table 1-19 for "The relationship between cognitive abilities and mental health as represented by cognitive abilities at the neural and genetic levels of analysis"

### SUPPLEMENTARY APPENDIX

SUPPLEMENTARY FIGURE 14. STACKED BAR PLOTS SHOWING COMMON AND UNIQUE EFFECTS OF PROXY MEASURES OF COGNITIVE ABILITIES BASED ON EACH SET OF NEUROIMAGING FEATURES IN EXPLAINING COGNITIVE ABILITIES ACROSS TEST SITES. 16

SUPPLEMENTARY TABLE 1. PERFORMANCE METRICS FOR PREDICTIVE MODELS, PREDICTING COGNITIVE ABILITIES FROM MENTAL HEALTH, NEUROIMAGING, POLYGENIC SCORES AND SOCIO-DEMOGRAPHICS, LIFESTYLES AND DEVELOPMENTS.22
Supplementary Table 10. Results of linear-mixed models using proxy measures of cognitive abilities based on mental health, neuroimaging, polygenic scores and/or socio-demographics, lifestyles and developmenta 31
Supplementary Table 11. Results of linear-mixed models using proxy measures of cognitive abilities based on mental health, neuroimaging, polygenic scores and/or socio-demographics, lifestyles and developmental adverse events as regressors to explain cognitive abilities across test sites in the follow-up. 32

#### Supplementary Figures

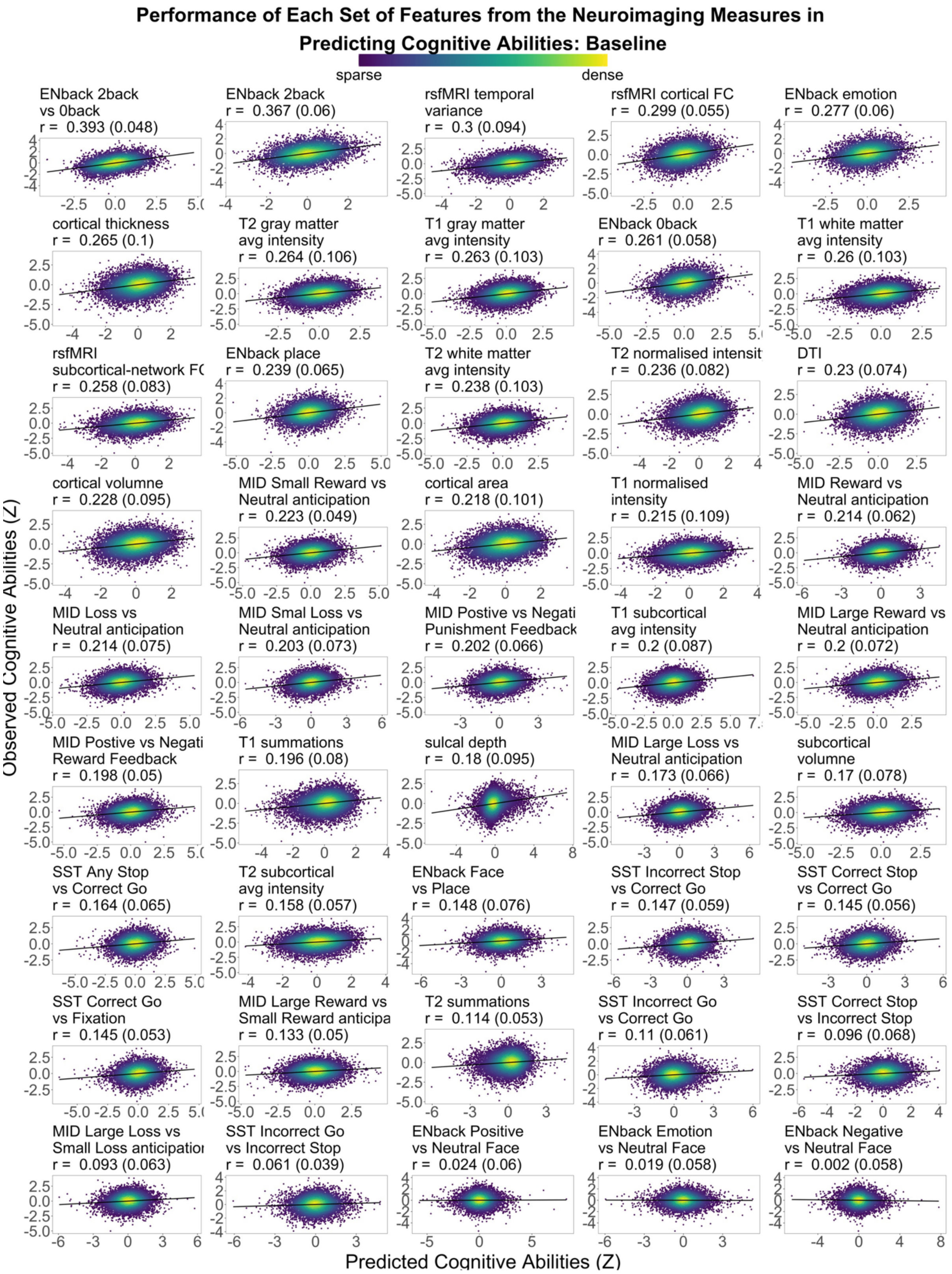

**Supplementary Figure 1. Scatter plots between observed vs predicted cognitive abilities based on each set of 45 neuroimaging features in the baseline data.** All data points are from test sets. *r* is the average Pearson's *r* across 21 test sites, and the parenthesis is the standard deviation of Pearson's *r* across sites.

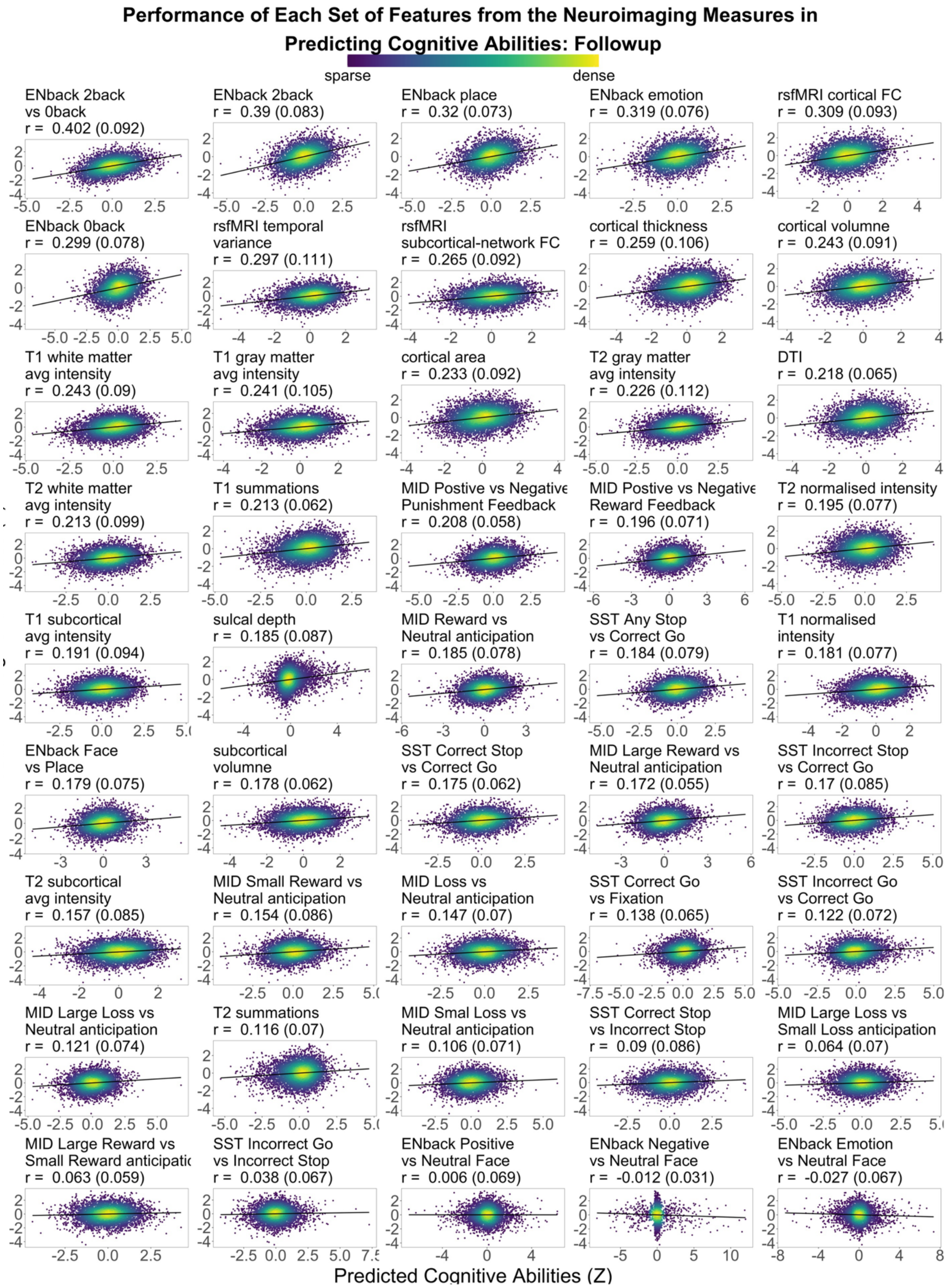

**Supplementary Figure 2. Scatter plots between observed vs predicted cognitive abilities based on each set of 45 neuroimaging features in the follow-up data.** All data points are from test sets. *r* is the average Pearson's *r* across 21 test sites, and the parenthesis is the standard deviation of Pearson's *r* across sites.

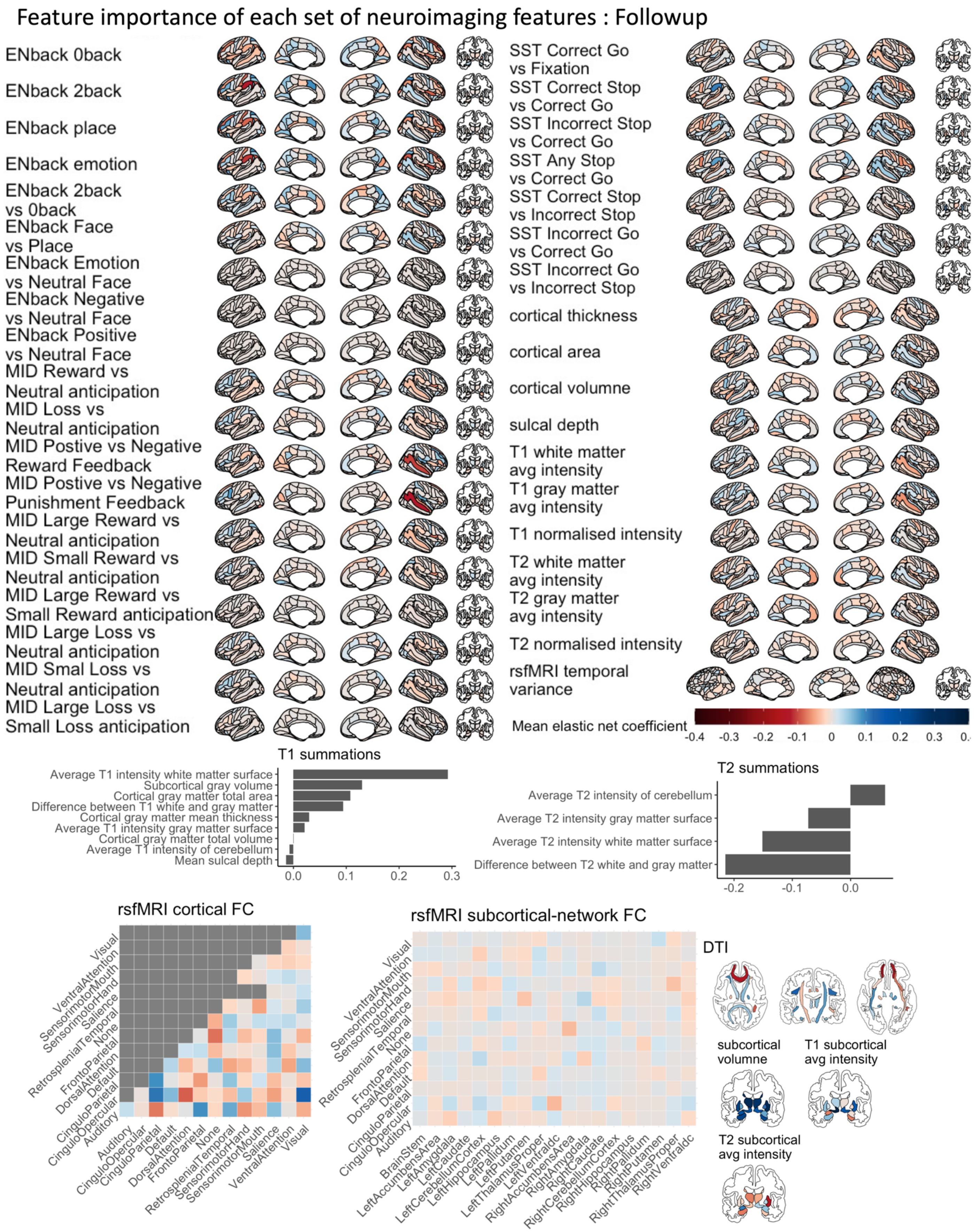

**Supplementary Figure 3. Feature importance of each set of neuroimaging features, predicting cognitive abilities in the follow-up data via Elastic Net.** The feature importance was based on the Elastic Net coefficient, averaged across test sites. We did not order these sets of neuroimaging features according to their feature importance (see Figure 2). MID = Monetary Incentive Delay task; SST = Stop Signal Task; DTI = Diffusion Tensor Imaging; FC = functional connectivity. The brain plots were created via the *ggseg* and *ggsegExtra* packages (1).

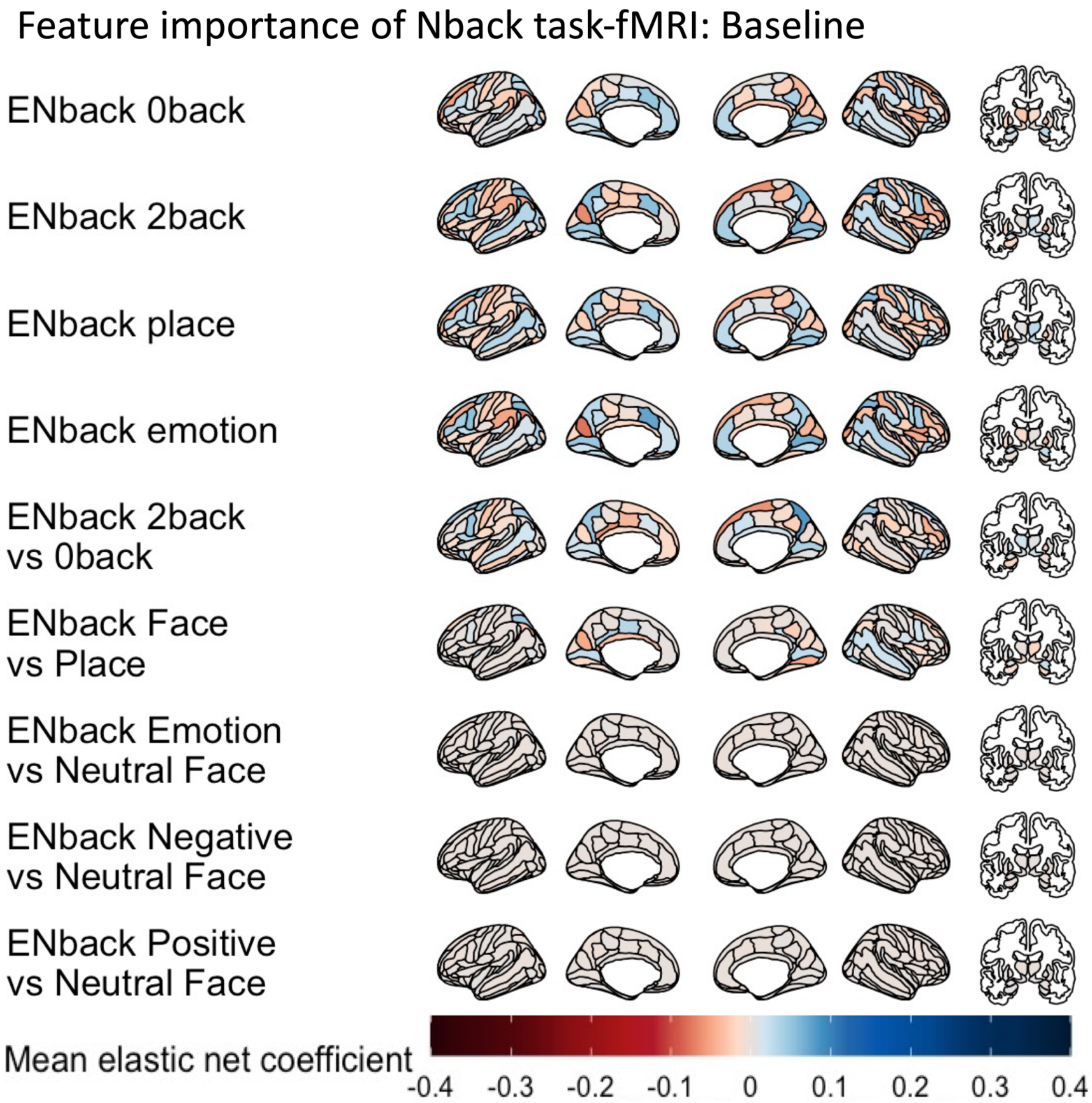

**Supplementary Figure 4. Feature importance of Nback task-fMRI features, predicting cognitive abilities in the baseline data via Elastic Net.** The feature importance was based on the Elastic Net coefficient, averaged across test sites. We did not order these sets of neuroimaging features according to their feature importance (see Figure 2). The brain plots were created via the *ggseg* and *ggsegExtra* packages (1).

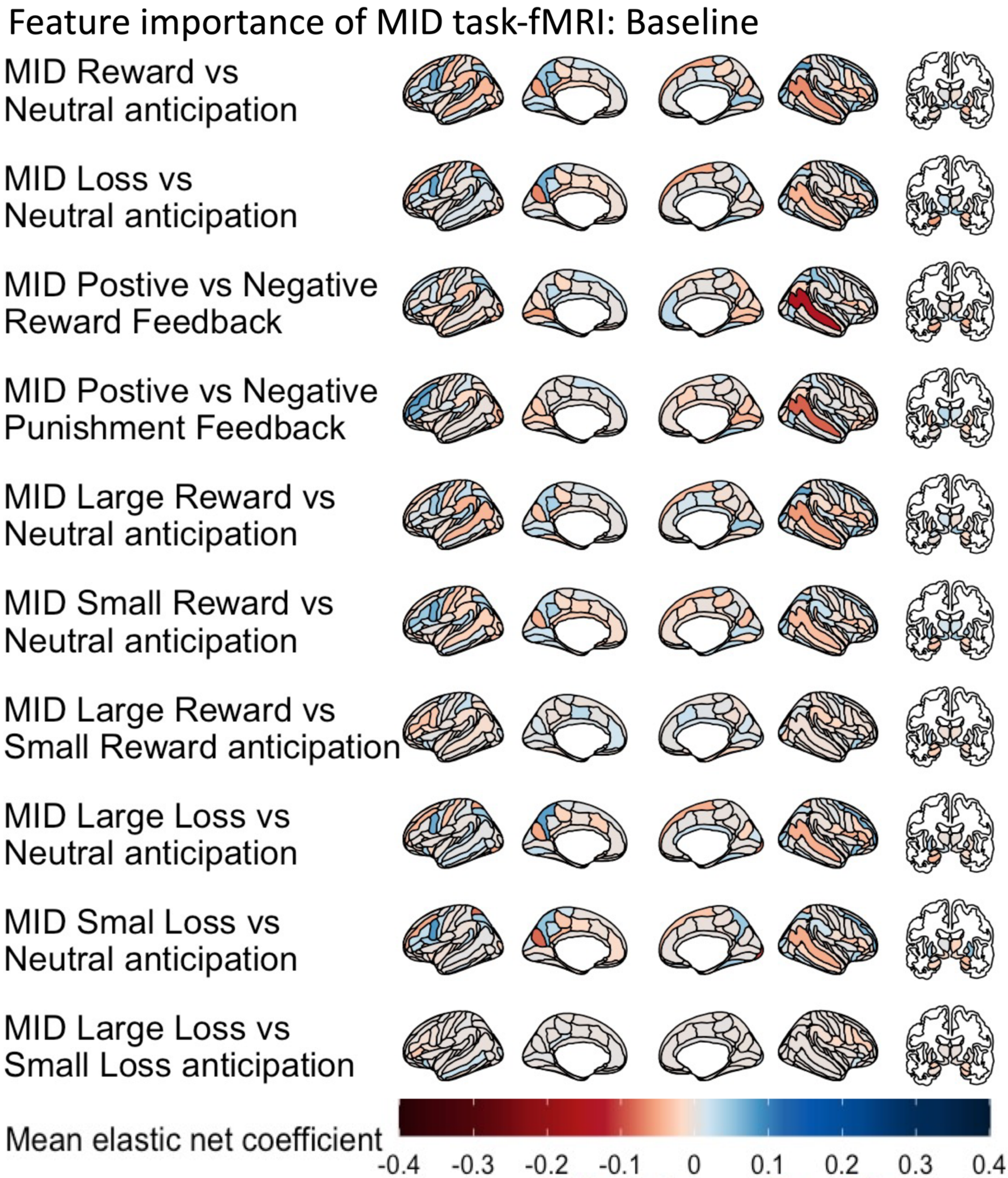

**Supplementary Figure 5. Feature importance of MID task-fMRI features, predicting cognitive abilities in the baseline data via Elastic Net.** The feature importance was based on the Elastic Net coefficient, averaged across test sites. We did not order these sets of neuroimaging features according to their feature importance (see Figure 2). MID = Monetary Incentive Delay task. The brain plots were created via the *ggseg* and *ggsegExtra* packages (1).

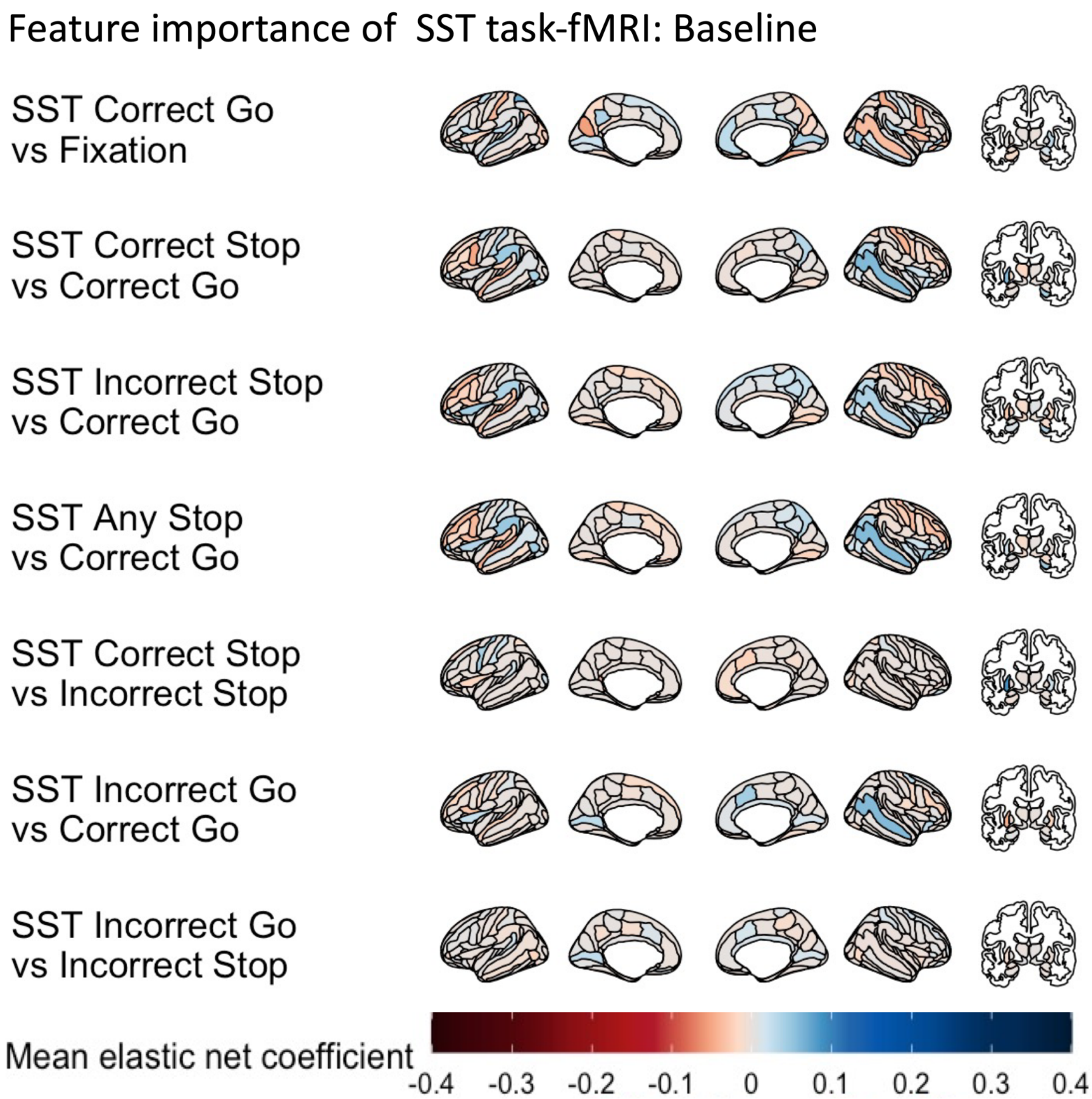

**Supplementary Figure 6. Feature importance of SST task-fMRI features, predicting cognitive abilities in the baseline data via Elastic Net.** The feature importance was based on the Elastic Net coefficient, averaged across test sites. We did not order these sets of neuroimaging features according to their feature importance (see Figure 2). SST = Stop Signal Task. The brain plots were created via the *ggseg* and *ggsegExtra* packages (1).

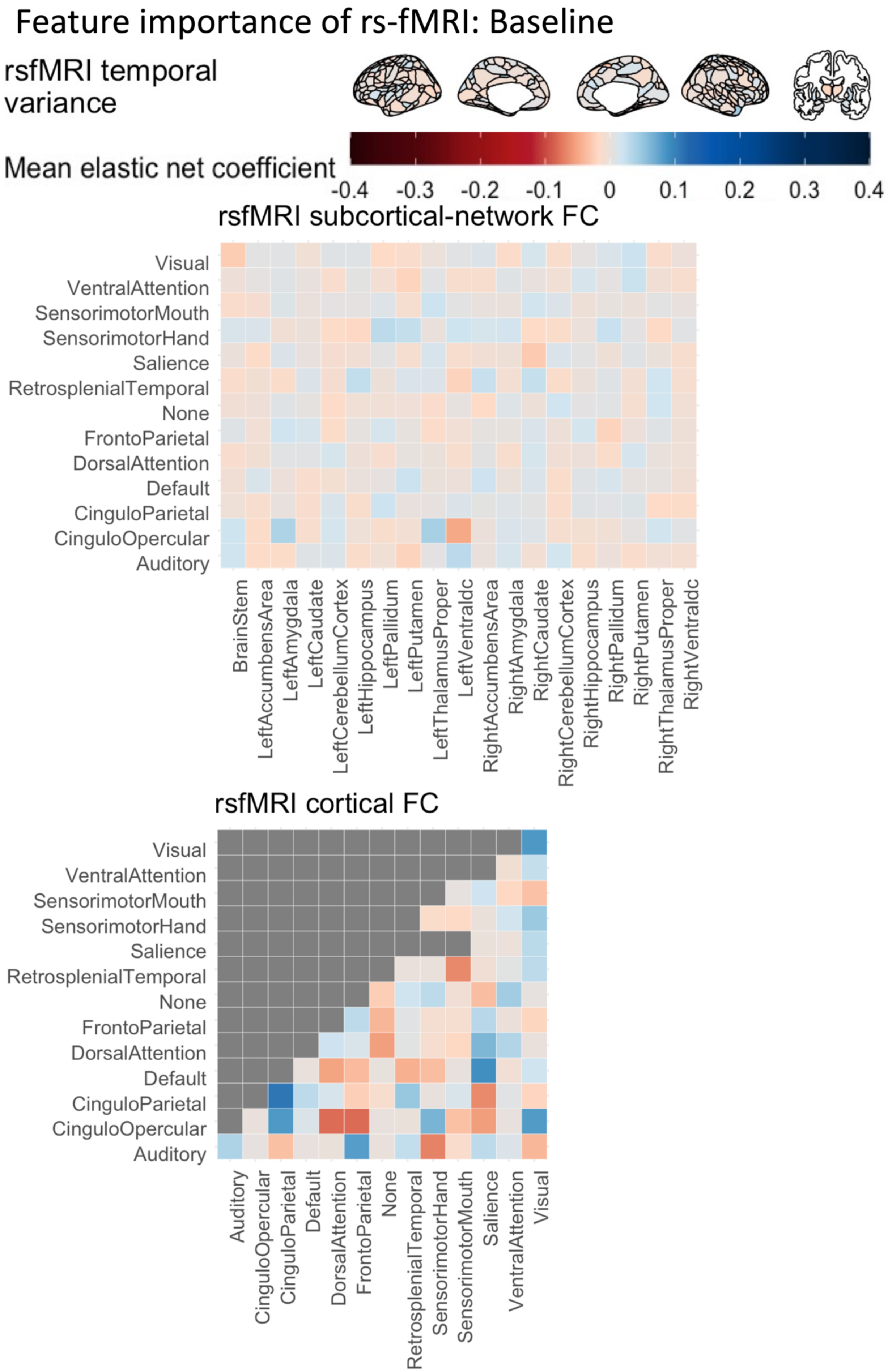

**Supplementary Figure 7. Feature importance of rs-fMRI features, predicting cognitive abilities in the baseline data via Elastic Net.** The feature importance was based on the Elastic Net coefficient, averaged across test sites. We did not order these sets of neuroimaging features according to their feature importance (see Figure 2). FC = functional connectivity. The brain plots were created via the *ggseg* and *ggsegExtra* packages (1).

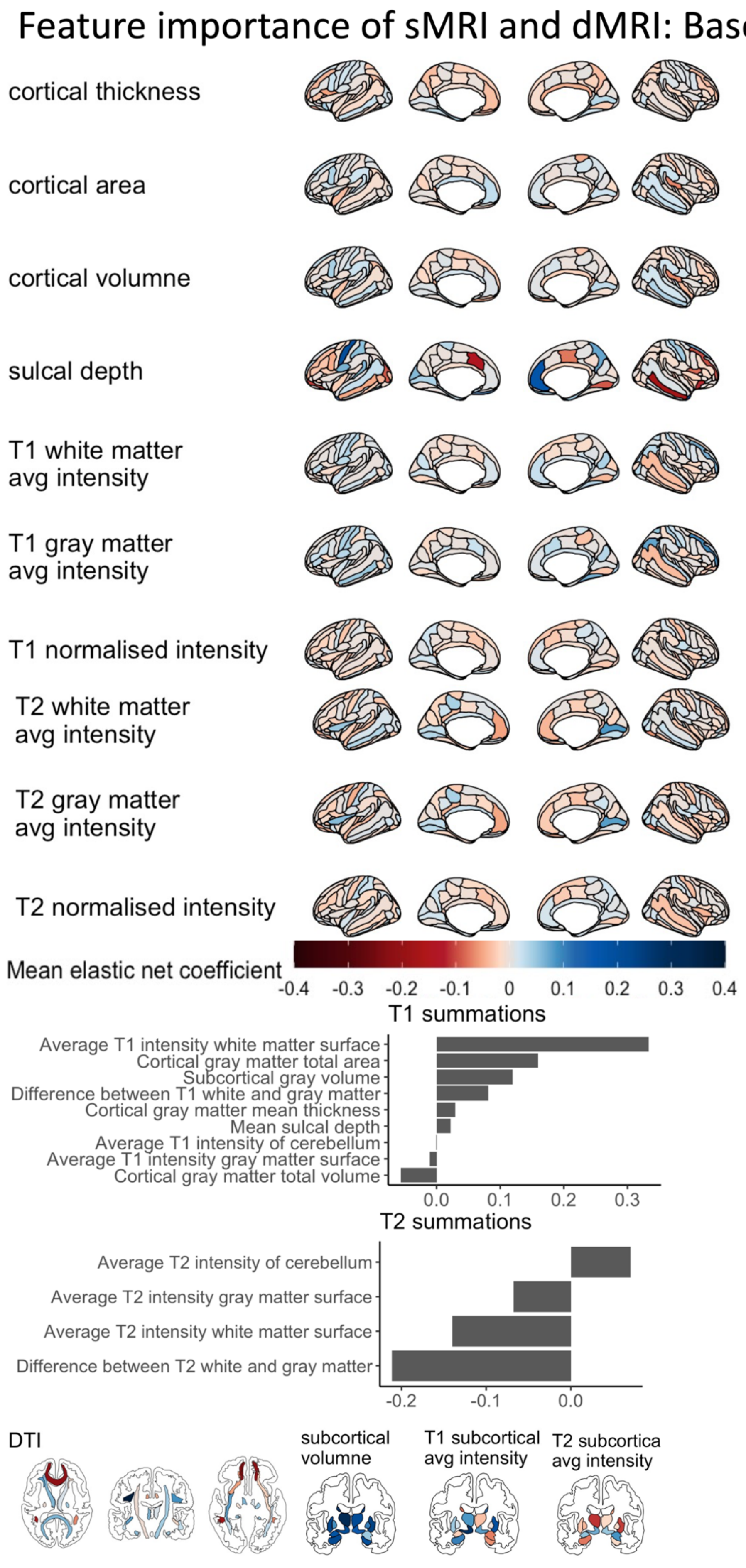

**Supplementary Figure 8. Feature importance of sMRI and dMRI features, predicting cognitive abilities in the baseline data via Elastic Net.** The feature importance was based on the Elastic Net coefficient, averaged across test sites. We did not order these sets of neuroimaging features according to their feature importance (see Figure 2). DTI = Diffusion Tensor Imaging. The brain plots were created via the *ggseg* and *ggsegExtra* packages (1).

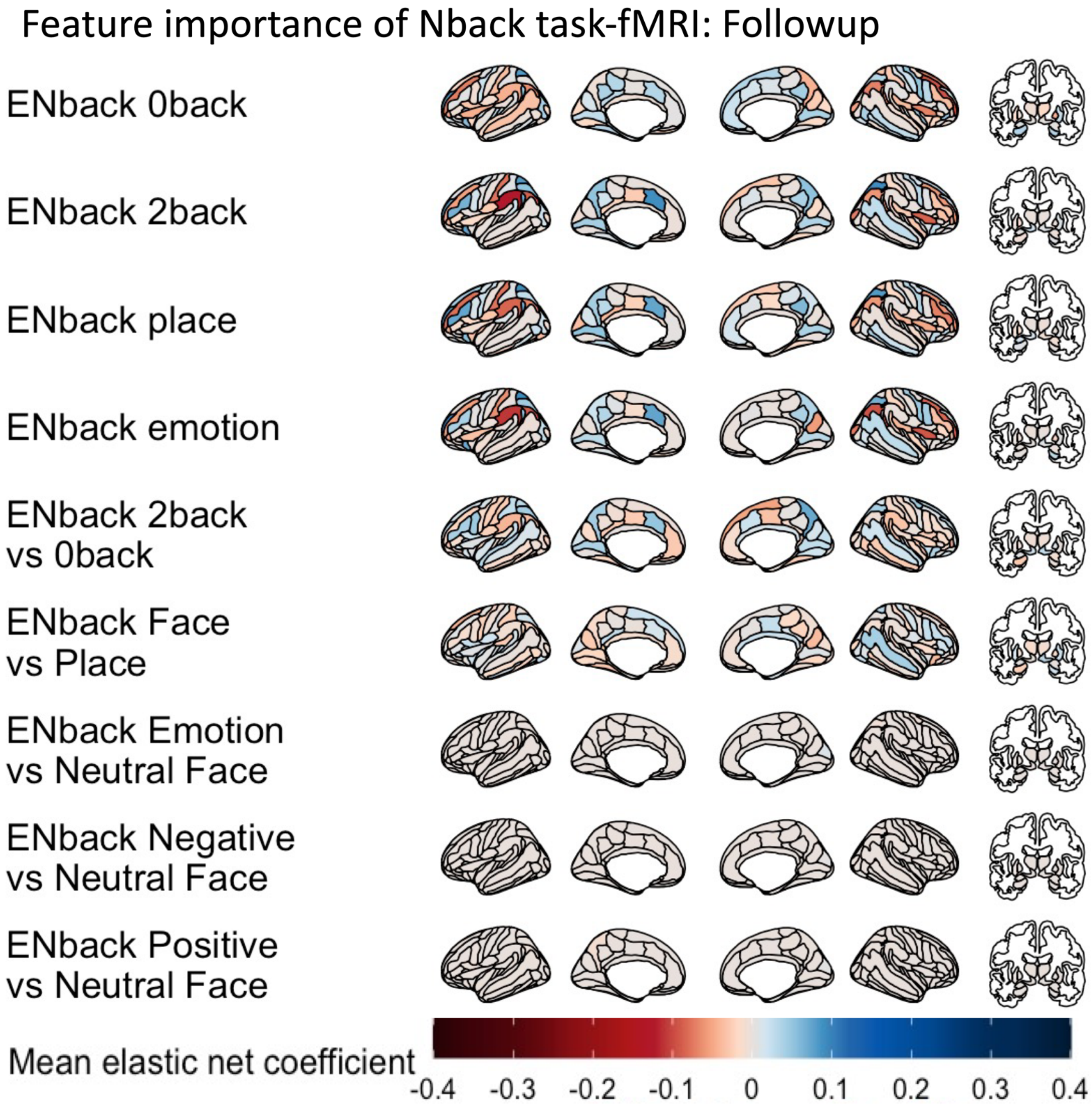

**Supplementary Figure 9. Feature importance of Nback task-fMRI features, predicting cognitive abilities in the follow-up data via Elastic Net.** The feature importance was based on the Elastic Net coefficient, averaged across test sites. We did not order these sets of neuroimaging features according to their feature importance (see Figure 2). The brain plots were created via the *ggseg* and *ggsegExtra* packages (1).

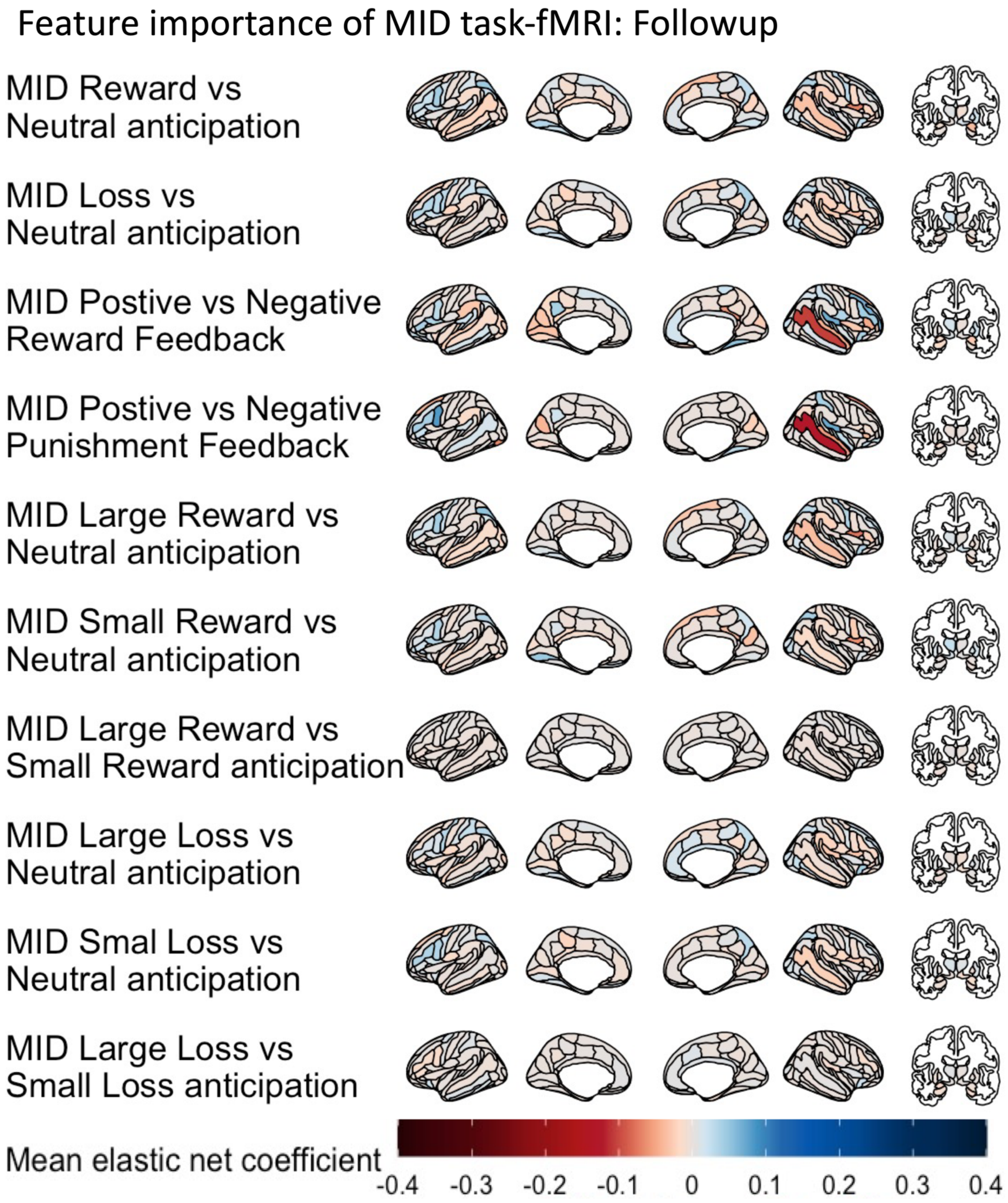

**Supplementary Figure 10. Feature importance of MID task-fMRI features, predicting cognitive abilities in the follow-up data via Elastic Net.** The feature importance was based on the Elastic Net coefficient, averaged across test sites. We did not order these sets of neuroimaging features according to their feature importance (see Figure 2). MID = Monetary Incentive Delay task. The brain plots were created via the *ggseg* and *ggsegExtra* packages (1).

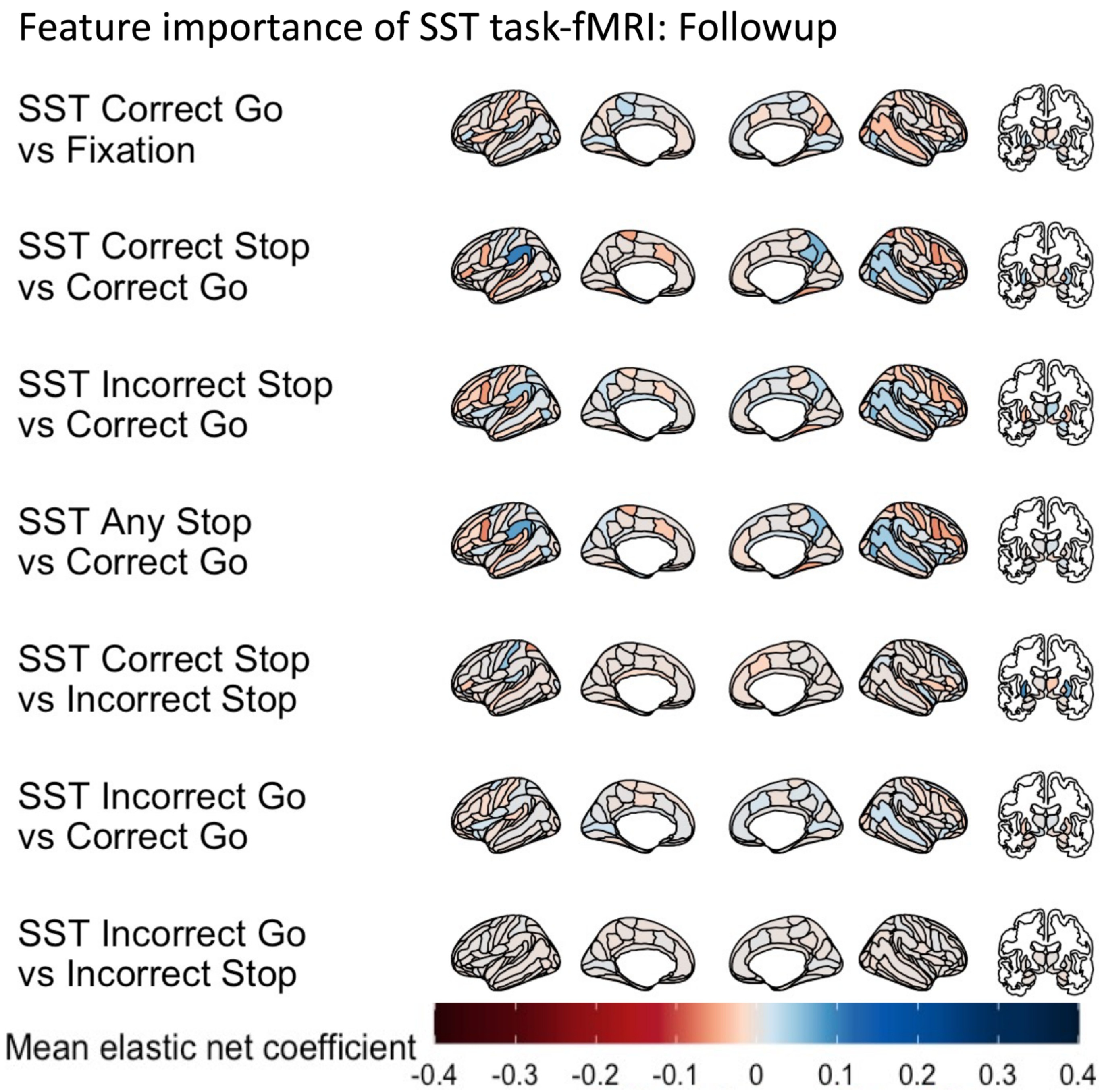

**Supplementary Figure 11. Feature importance of SST task-fMRI features, predicting cognitive abilities in the follow-up data via Elastic Net.** The feature importance was based on the Elastic Net coefficient, averaged across test sites. We did not order these sets of neuroimaging features according to their feature importance (see Figure 2). SST = Stop Signal Task. The brain plots were created via the *ggseg* and *ggsegExtra* packages (1).

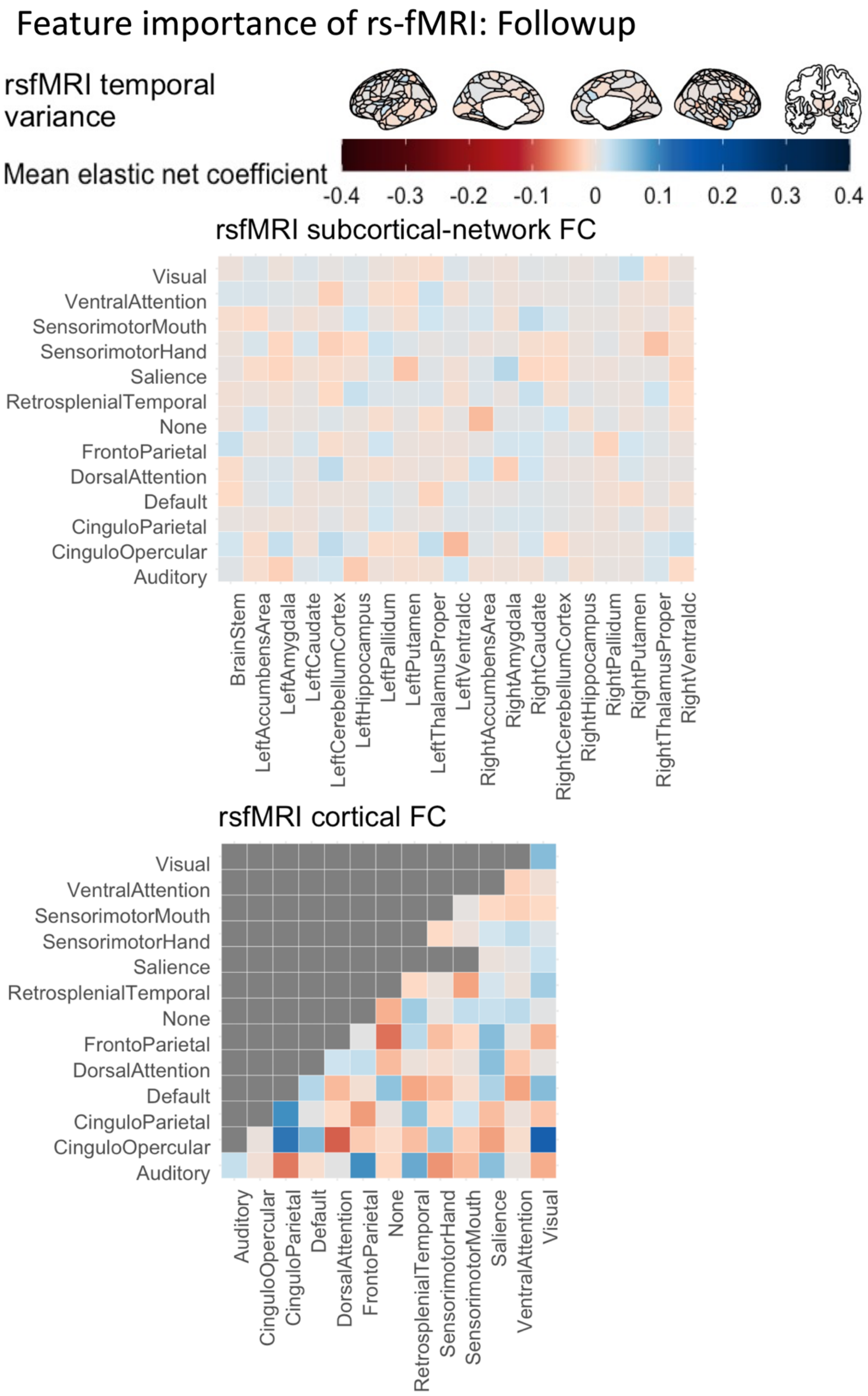

**Supplementary Figure 12. Feature importance of rs-fMRI features, predicting cognitive abilities in the follow-up data via Elastic Net.** The feature importance was based on the Elastic Net coefficient, averaged across test sites. We did not order these sets of neuroimaging features according to their feature importance (see Figure 2). FC = functional connectivity. The brain plots were created via the *ggseg* and *ggsegExtra* packages (1).

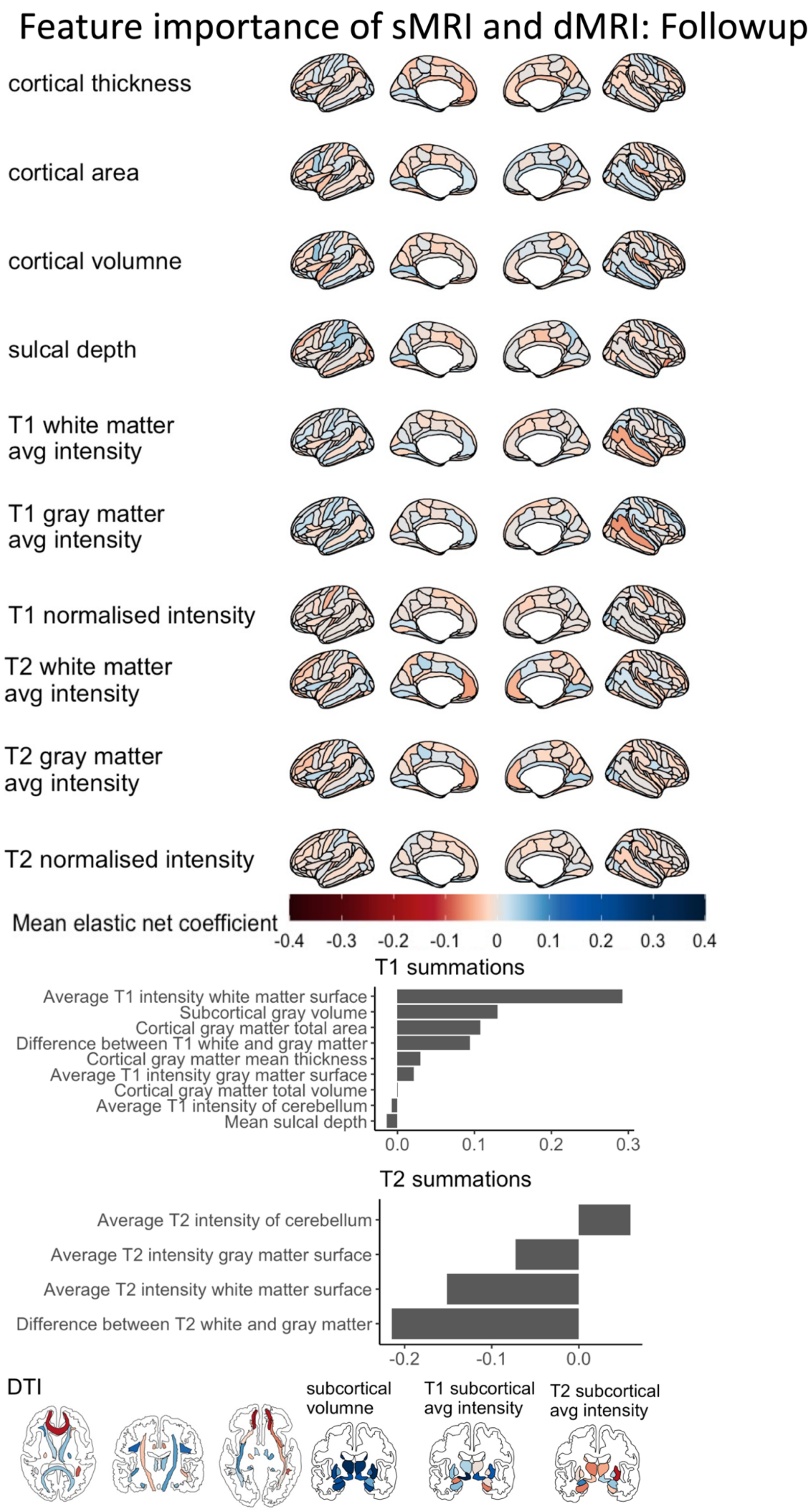

**Supplementary Figure 13. Feature importance of sMRI and dMRI features, predicting cognitive abilities in the follow-up data via Elastic Net.** The feature importance was based on the Elastic Net coefficient, averaged across test sites. We did not order these sets of neuroimaging features according to their feature importance (see Figure 2). DTI = Diffusion Tensor Imaging. The brain plots were created via the *ggseg* and *ggsegExtra* packages (1).

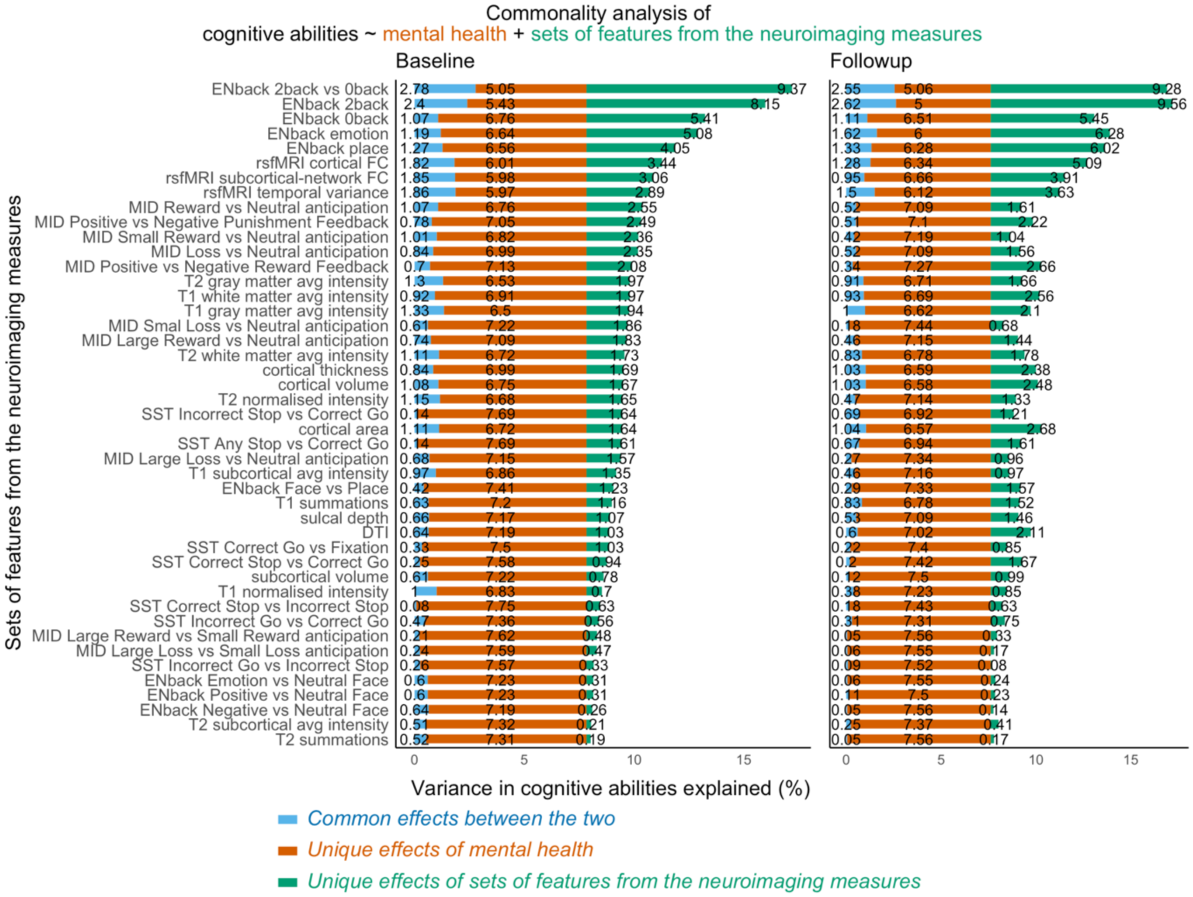

**Supplementary Figure 14. Stacked bar plots showing common and unique effects of proxy measures of cognitive abilities based on each set of neuroimaging features in explaining cognitive abilities across test sites.** We computed the common and unique effects in % based on the marginal $R^2$ of linear-mixed models.

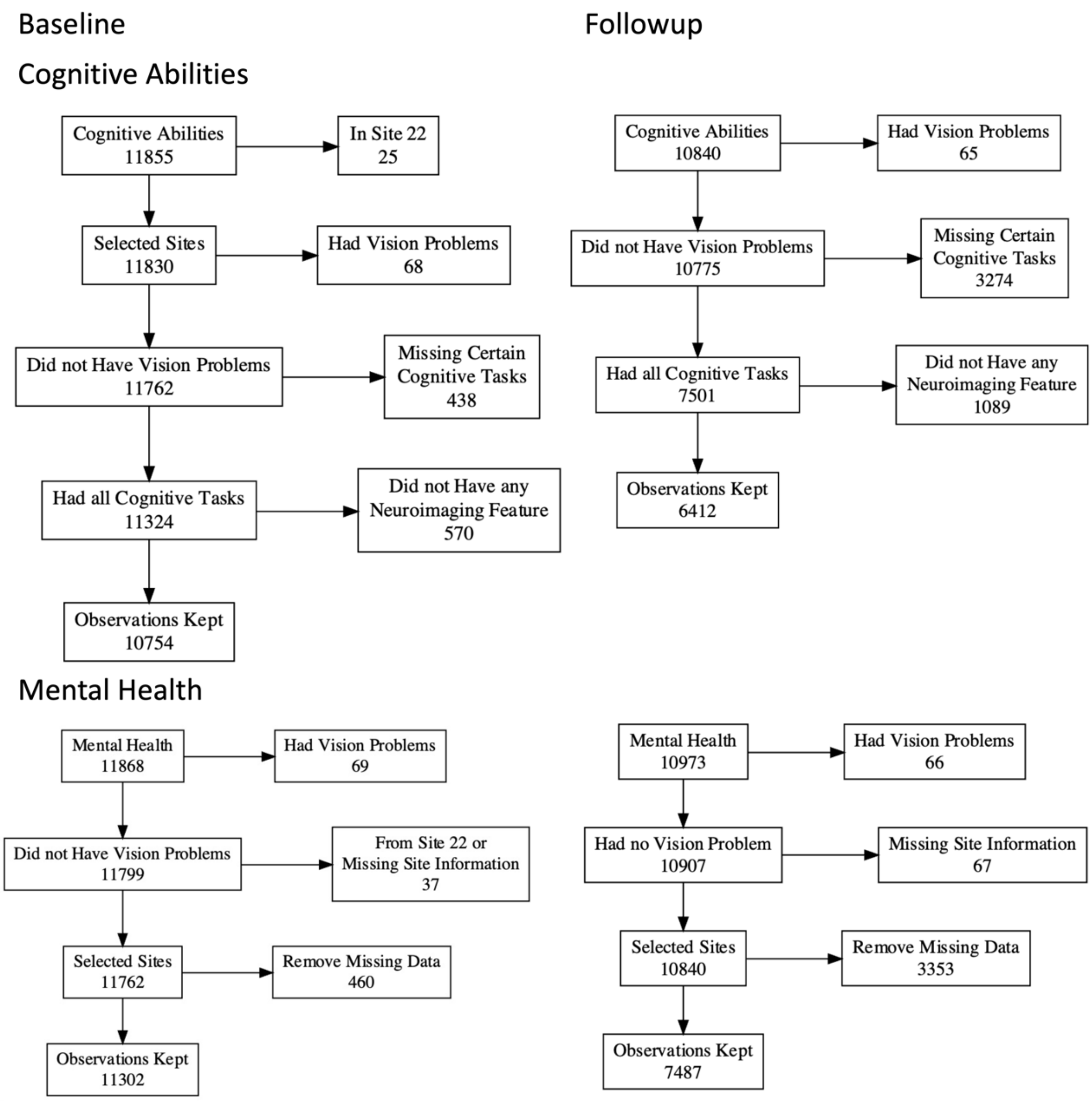

**Supplementary Figure 15. Flow diagram of participants' inclusion and exclusion criteria.** Here we show the criteria for cognitive abilities and mental health across the two time points.

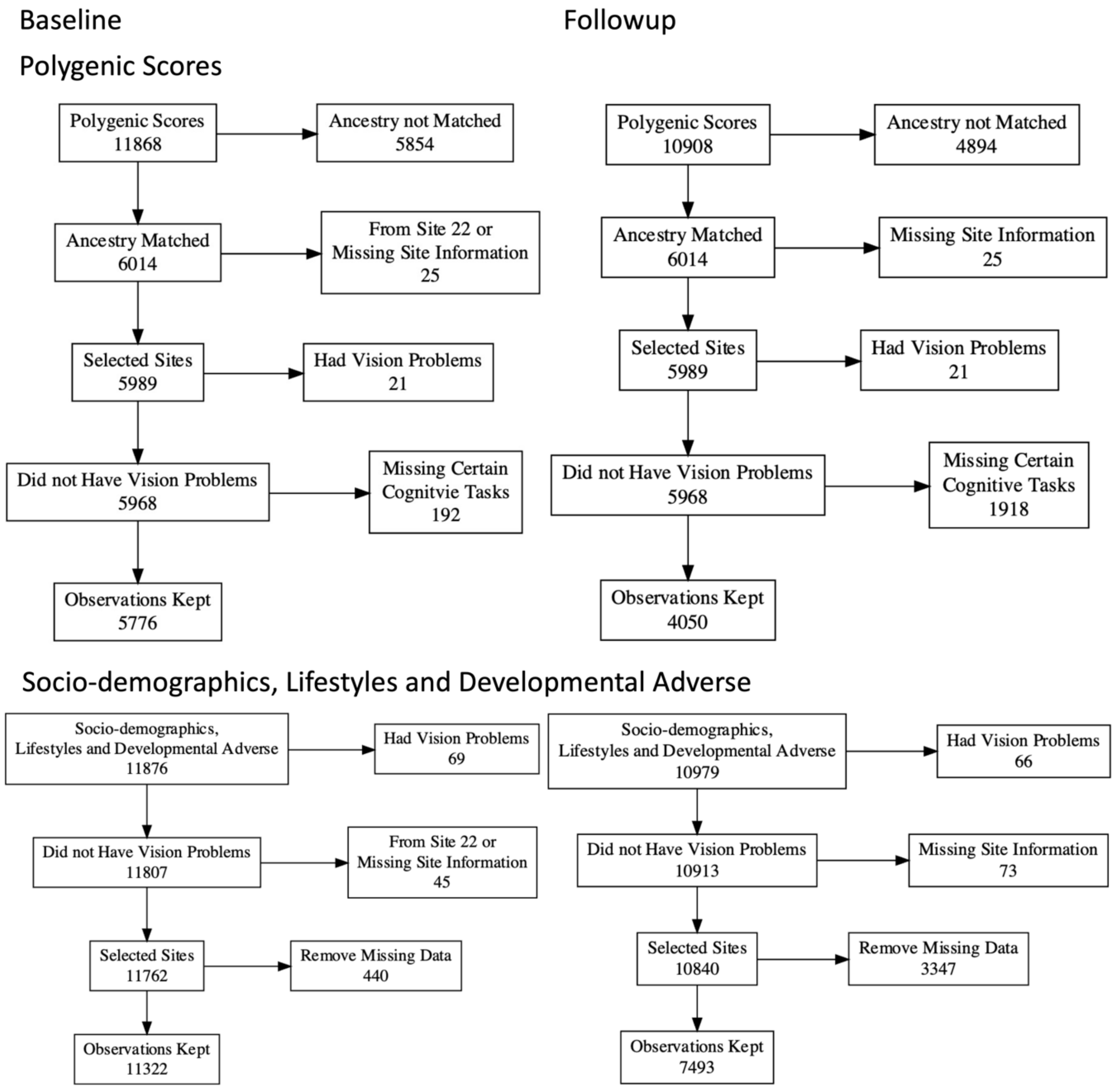

**Supplementary Figure 15 (Cont.). Flow diagram of participants' inclusion and exclusion criteria.** Here we show the criteria for polygenic scores and social demographics, lifestyle and developmental adverse events across the two time points.

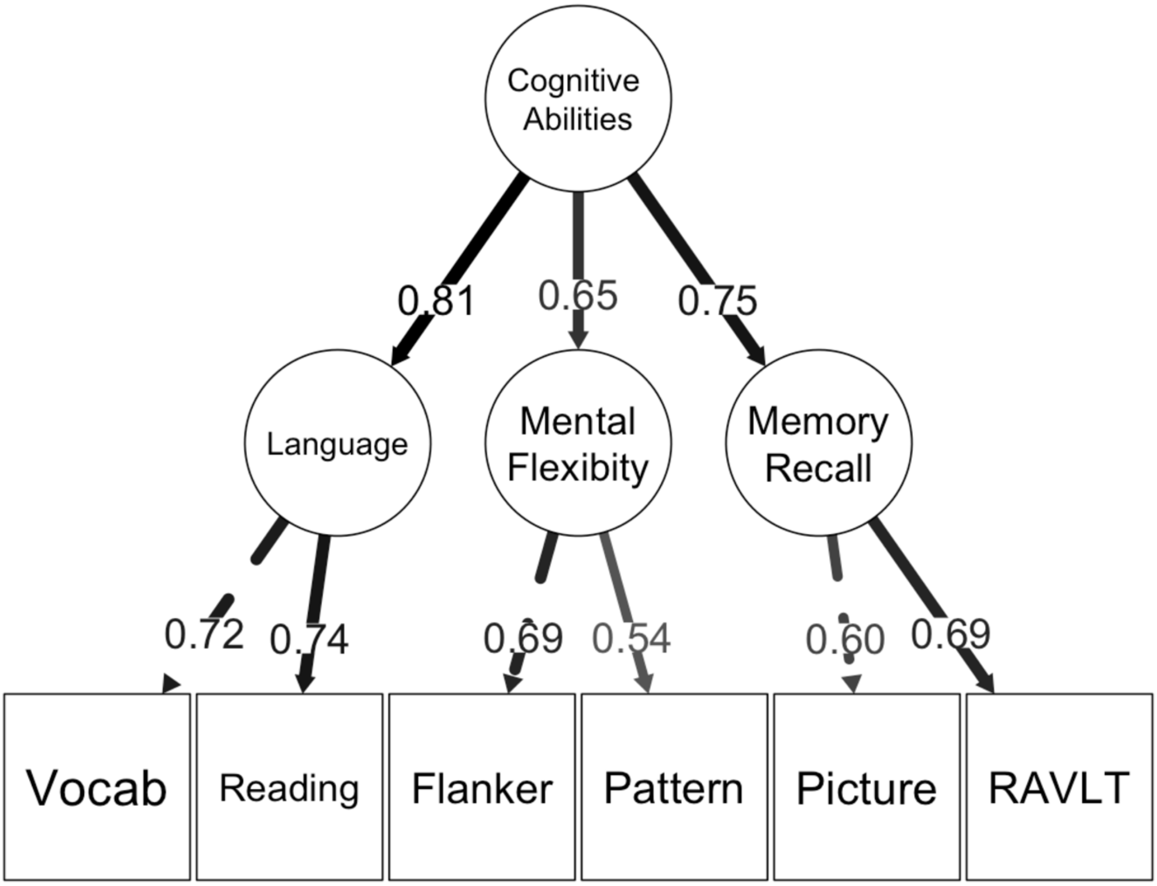

**Supplementary Figure 16. Standardised weights of the second-order 'g-factor' model.** These weights were derived from confirmatory factor analysis, fitted on cognitive abilities across six cognitive tasks from the entire baseline dataset. The actual weights used for predictive modelling were slightly different, as the predictive modelling was based on leave-one-site-out cross-validation, which trained on data from all but one site.

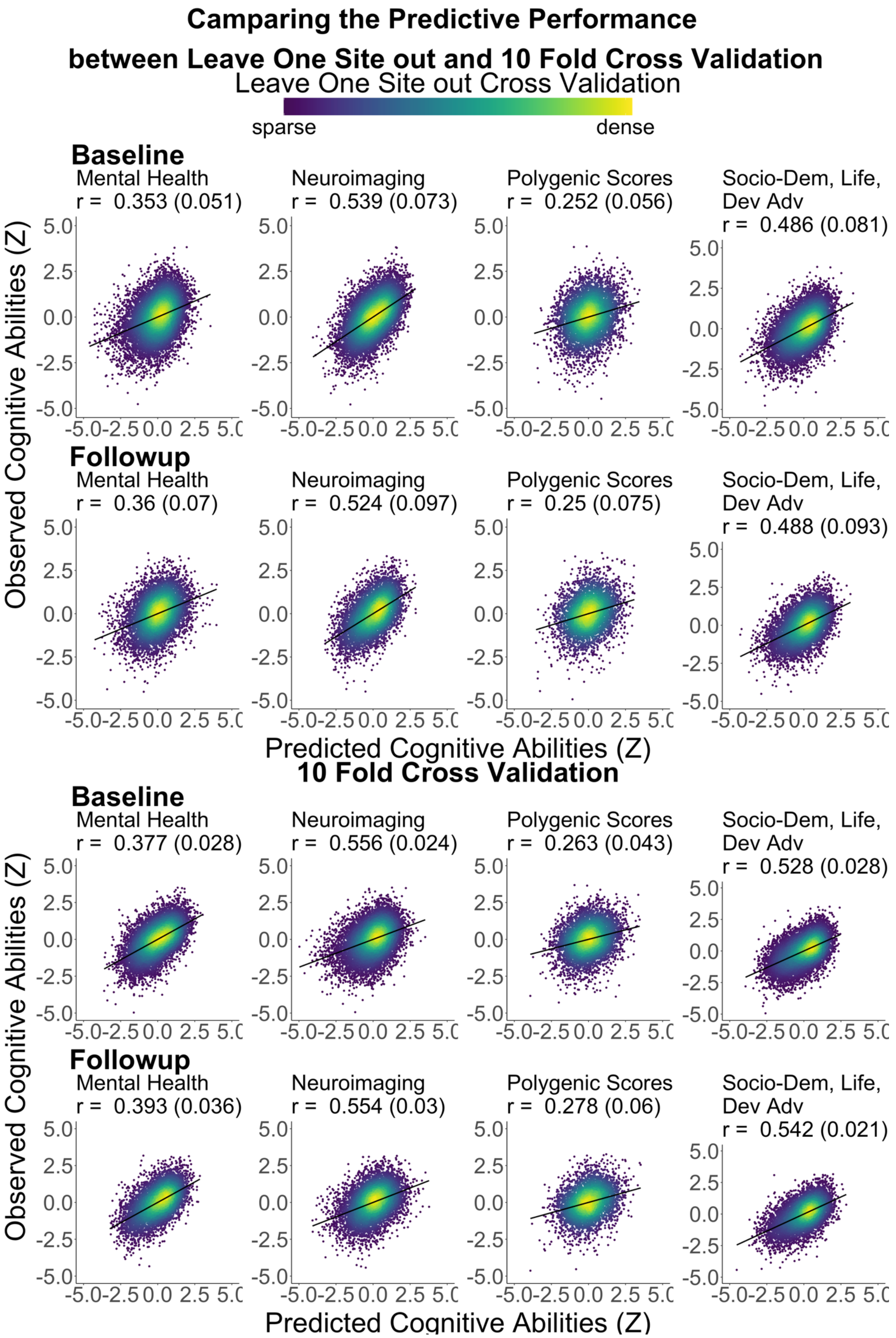

**Supplementary Figure 17. Predictive performance of leave one site out cross validation vs tenfold cross validation.**

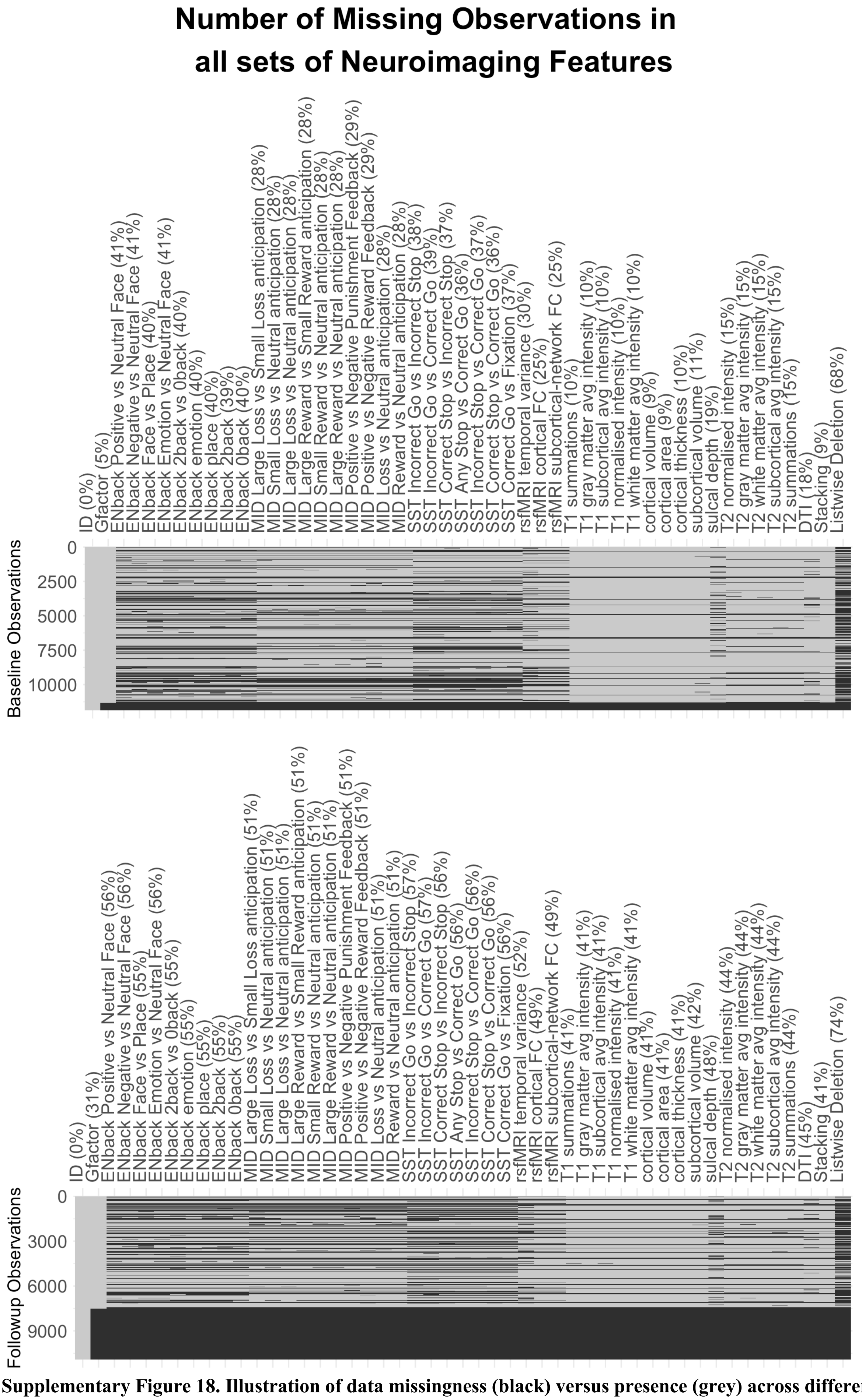

**Supplementary Figure 18. Illustration of data missingness (black) versus presence (grey) across different sets of neuroimaging features.** This figure compares the number of observations in the analysis. Opportunist stacking (referred to as stacking here) requires only at least one neuroimaging feature to be present, thus allowing the inclusion of more neuroimaging features compared to listwise deletion.

#### Supplementary Tables

**Supplementary Table 1. Performance metrics for predictive models, predicting cognitive abilities from mental health, neuroimaging, polygenic scores and socio-demographics, lifestyles and developments.** The metrics were averaged across test sites with standard deviations in parentheses. $R^2$ = coefficient of determination; MAE = mean-absolute error; RMSE = root mean square error.

| Features | Correlation | $R^2$ | MAE | RMSE |
|---|---|---|---|---|
| Baseline | | | | |
| Mental Health | 0.353 (0.051) | 0.124 (0.038) | 0.736 (0.019) | 0.934(0.02) |
| CBCL | 0.272 (0.048) | 0.074 (0.028) | 0.758 (0.014) | 0.961 (0.015) |
| Child personality | 0.268 (0.058) | 0.071 (0.034) | 0.759 (0.019) | 0.962 (0.017) |
| Neuroimaging | 0.539 (0.073) | 0.291 (0.082) | 0.658 (0.039) | 0.839 (0.05) |
| Polygenic scores | 0.252 (0.056) | 0.02 (0.075) | 0.696 (0.055) | 0.884 (0.066) |
| Socio-demo Life Dev Adv | 0.486 (0.081) | 0.239 (0.084) | 0.686 (0.041) | 0.87 (0.049) |
| Follow-up | | | | |
| Mental Health | 0.36 (0.07) | 0.116 (0.061) | 0.715 (0.043) | 0.903 (0.051) |
| CBCL | 0.24 (0.056) | 0.043 (0.034) | 0.746 (0.045) | 0.94 (0.053) |
| Child personality | 0.311 (0.076) | 0.084 (0.059) | 0.728 (0.046) | 0.919 (0.051) |
| Neuroimaging | 0.524 (0.097) | 0.266 (0.112) | 0.645 (0.038) | 0.818 (0.053) |
| Polygenic scores | 0.25 (0.075) | 0.031 (0.068) | 0.672 (0.053) | 0.854 (0.068) |
| Socio-demo Life Dev Adv | 0.488 (0.093) | 0.226 (0.096) | 0.664 (0.044) | 0.843 (0.05) |

**Supplementary Table 2. Performance metrics for predictive models, predicting cognitive abilities from the 45 sets of neuroimaging features in the baseline data.** The metrics were averaged across test sites with standard deviations in parentheses. $R^2$ = coefficient of determination; MAE = mean-absolute error; RMSE = root mean square error.

| Features | Correlation | $R^2$ | MAE | RMSE |
|---|---|---|---|---|
| Neuroimaging | 0.539 (0.073) | 0.291 (0.082) | 0.658 (0.039) | 0.839 (0.05) |
| ENback 2back vs 0back | 0.393 (0.048) | 0.147 (0.042) | 0.661 (0.038) | 0.841 (0.045) |
| ENback 2back | 0.367 (0.06) | 0.128 (0.048) | 0.667 (0.036) | 0.848 (0.043) |
| rsfMRI temporal variance | 0.3 (0.094) | 0.09 (0.054) | 0.728 (0.04) | 0.921 (0.045) |
| rsfMRI cortical FC | 0.299 (0.055) | 0.088 (0.034) | 0.734 (0.027) | 0.929 (0.032) |
| ENback emotion | 0.277 (0.06) | 0.07 (0.041) | 0.689 (0.031) | 0.876 (0.035) |
| cortical thickness | 0.265 (0.1) | 0.072 (0.055) | 0.756 (0.026) | 0.96 (0.03) |
| T2 gray matter avg intensity | 0.264 (0.106) | 0.069 (0.064) | 0.752 (0.032) | 0.953 (0.035) |
| T1 gray matter avg intensity | 0.263 (0.103) | 0.063 (0.071) | 0.761 (0.033) | 0.965 (0.039) |
| ENback 0back | 0.261 (0.058) | 0.061 (0.038) | 0.688 (0.031) | 0.878 (0.035) |
| T1 white matter avg intensity | 0.26 (0.103) | 0.067 (0.063) | 0.76 (0.029 | 0.963 (0.035) |
| rsfMRI subcortical-network FC | 0.258 (0.083) | 0.066 (0.043) | 0.743 (0.033) | 0.94 (0.035) |
| ENback place | 0.239 (0.065) | 0.049 (0.041) | 0.695 (0.032) | 0.886 (0.038) |
| T2 white matter avg intensity | 0.238 (0.103 | 0.056 (0.056) | 0.756 (0.03) | 0.96 (0.031) |
| T2 normalised intensity | 0.236 (0.082) | 0.057 (0.041) | 0.755 (0.021) | 0.96 (0.024) |
| DTI | 0.23 (0.074) | 0.042 (0.048) | 0.762 (0.027) | 0.967 (0.029) |
| cortical volume | 0.228 (0.095) | 0.053 (0.044) | 0.767 (0.02) | 0.971 (0.024) |
| MID Small Rew vs Neu anticipation | 0.223 (0.049) | 0.048 (0.022) | 0.743 (0.017) | 0.938 (0.02) |
| cortical area | 0.218 (0.101) | 0.049 (0.046) | 0.768 (0.021) | 0.973 (0.025) |
| T1 normalised intensity | 0.215 (0.109) | 0.047 (0.049) | 0.769 (0.022) | 0.974 (0.028) |
| MID Reward vs Neutral anticipation | 0.214 (0.062) | 0.043 (0.028) | 0.745 (0.022) | 0.944 (0.024) |
| MID Loss vs Neutral anticipation | 0.214 (0.075) | 0.043 (0.034) | 0.745 (0.025) | 0.944 (0.028) |
| MID Small Loss vs Neu anticipation | 0.203 (0.073) | 0.038 (0.03) | 0.747 (0.026) | 0.945 (0.026) |
| MID Pos vs Neg Punishment Feedback | 0.202 (0.066) | 0.037 (0.027) | 0.745 (0.021) | 0.945 (0.026) |
| T1 subcortical avg intensity | 0.2 (0.087) | 0.037 (0.043) | 0.773 (0.023) | 0.979 (0.026) |
| MID Large Rew vs Neu anticipation | 0.2 (0.072) | 0.037 (0.03) | 0.747 (0.021) | 0.946 (0.024) |
| MID Pos vs Neg Reward Feedback | 0.198 (0.05) | 0.036 (0.02) | 0.748 (0.022) | 0.945 (0.028) |
| T1 summations | 0.196 (0.08) | 0.009 (0.059) | 0.784 (0.029) | 0.992 (0.033) |
| sulcal depth | 0.18 (0.095) | 0.032 (0.039) | 0.777 (0.02) | 0.984 (0.026) |
| MID Large Loss vs Neu anticipation | 0.173 (0.066) | 0.026 (0.026) | 0.749 (0.022) | 0.95 (0.025) |
| subcortical volume | 0.17 (0.078) | 0.028 (0.029) | 0.775 (0.018) | 0.982 (0.021) |
| SST Any Stop vs Correct Go | 0.164 (0.065) | 0.022 (0.025) | 0.736 (0.038) | 0.935 (0.043) |
| T2 subcortical avg intensity | 0.158 (0.057) | 0.023 (0.023) | 0.77 (0.018) | 0.977 (0.02) |
| ENback Face vs Place | 0.148 (0.076) | 0.014 (0.028) | 0.712 (0.027) | 0.904 (0.034) |
| SST Incorrect Stop vs Correct Go | 0.147 (0.059) | 0.017 (0.02) | 0.738 (0.035) | 0.937 (0.04) |
| SST Correct Stop vs Correct Go | 0.145 (0.056) | 0.017 (0.018) | 0.739 (0.033) | 0.936 (0.038) |
| SST Correct Go vs Fixation | 0.145 (0.053) | 0.017 (0.017) | 0.74 (0.033) | 0.938 (0.036) |
| MID Large Rew vs Small anticipation | 0.133 (0.05) | 0.015 (0.014) | 0.757 (0.022) | 0.956 (0.025) |
| T2 summations | 0.114 (0.053) | 0.008 (0.022) | 0.777 (0.018) | 0.984 (0.016) |
| SST Incorrect Go vs Correct Go | 0.11 (0.061) | 0.008 (0.015) | 0.744 (0.034) | 0.94 (0.038) |
| SST Correct Stop vs Incorrect Stop | 0.096 (0.068) | 0.005 (0.018) | 0.744 (0.033) | 0.943 (0.036) |
| MID Large vs Small Loss anticipation | 0.093 (0.063) | 0.006 (0.014) | 0.756 (0.024) | 0.96 (0.026) |
| SST Incorrect Go vs Incorrect Stop | 0.061 (0.039) | 0 (0.008) | 0.744 (0.032) | 0.943 (0.036) |
| ENback Positive vs Neutral Face | 0.024 (0.06) | -0.007 (0.012) | 0.716 (0.027) | 0.908 (0.034) |
| ENback Emotion vs Neutral Face | 0.019 (0.058) | -0.007 (0.01) | 0.716 (0.026) | 0.908 (0.033) |
| ENback Negative vs Neutral Face | 0.002 (0.058) | -0.007 (0.009) | 0.718 (0.024) | 0.911 (0.03) |

**Supplementary Table 3. Performance metrics for predictive models, predicting cognitive abilities from the 45 sets of neuroimaging features in the follow-up data.** The metrics were averaged across test sites with standard deviations in parentheses. $R^2$ = coefficient of determination; MAE = mean-absolute error; RMSE = root mean square error.

| Features | Correlation | $R^2$ | MAE | RMSE |
|---|---|---|---|---|
| Neuroimaging | 0.524 (0.097) | 0.266 (0.112) | 0.645 (0.038) | 0.818 (0.053) |
| ENback 2back vs 0back | 0.402 (0.092) | 0.15 (0.075) | 0.671 (0.032) | 0.844 (0.041) |
| ENback 2back | 0.39 (0.083) | 0.14 (0.071) | 0.676 (0.036) | 0.848 (0.045) |
| ENback place | 0.32 (0.073) | 0.089 (0.049) | 0.695 (0.038) | 0.874 (0.047) |
| ENback emotion | 0.319 (0.076) | 0.089 (0.05) | 0.696 (0.04) | 0.876 (0.047) |
| rsfMRI cortical FC | 0.309 (0.093) | 0.081 (0.071) | 0.718 (0.037) | 0.908 (0.046) |
| ENback 0back | 0.299 (0.078) | 0.077 (0.057) | 0.7 (0.045) | 0.881 (0.052) |
| rsfMRI temporal variance | 0.297 (0.111) | 0.077 (0.071) | 0.718 (0.045) | 0.903 (0.052) |
| rsfMRI subcortical-network FC | 0.265 (0.092) | 0.056 (0.059) | 0.732 (0.039) | 0.92 (0.048) |
| cortical thickness | 0.259 (0.106) | 0.055 (0.062) | 0.738 (0.034) | 0.932 (0.041) |
| cortical volume | 0.243 (0.091) | 0.046 (0.049) | 0.744 (0.034) | 0.936 (0.039) |
| T1 white matter avg intensity | 0.243 (0.09) | 0.044 (0.057) | 0.742 (0.035) | 0.937 (0.042) |
| T1 gray matter avg intensity | 0.241 (0.105) | 0.04 (0.069) | 0.742 (0.039) | 0.939 (0.047) |
| cortical area | 0.233 (0.092) | 0.041 (0.05) | 0.746 (0.032) | 0.939 (0.04) |
| T2 gray matter avg intensity | 0.226 (0.112) | 0.04 (0.064) | 0.743 (0.037) | 0.939 (0.049) |
| DTI | 0.218 (0.065) | 0.022 (0.052) | 0.747 (0.034) | 0.944 (0.041) |
| T2 white matter avg intensity | 0.213 (0.099) | 0.033 (0.057) | 0.747 (0.036) | 0.942 (0.045) |
| T1 summations | 0.213 (0.062) | 0.011 (0.046) | 0.756 (0.039) | 0.954 (0.044) |
| MID Pos vs Neg Punish Feedback | 0.208 (0.058) | 0.025 (0.033) | 0.743 (0.044) | 0.933 (0.049) |
| MID Pos vs Neg Reward Feedback | 0.196 (0.071) | 0.021 (0.042) | 0.742 (0.038) | 0.933 (0.042) |
| T2 normalised intensity | 0.195 (0.077) | 0.025 (0.035) | 0.749 (0.039) | 0.946 (0.045) |
| T1 subcortical avg intensity | 0.191 (0.094) | 0.002 (0.083) | 0.759 (0.039) | 0.957 (0.046) |
| sulcal depth | 0.185 (0.087) | 0.018 (0.048) | 0.756 (0.034) | 0.95 (0.043) |
| MID Reward vs Neutral anticipation | 0.185 (0.078) | 0.016 (0.039) | 0.746 (0.037) | 0.937 (0.04) |
| SST Any Stop vs Correct Go | 0.184 (0.079) | 0.018 (0.034) | 0.745 (0.047) | 0.934 (0.054) |
| T1 normalised intensity | 0.181 (0.077) | 0.018 (0.036) | 0.752 (0.038) | 0.95 (0.045) |
| ENback Face vs Place | 0.179 (0.075) | 0.019 (0.03) | 0.721 (0.039) | 0.907 (0.044) |
| subcortical volume | 0.178 (0.062) | 0.016 (0.032) | 0.752 (0.036) | 0.949 (0.041) |
| SST Correct Stop vs Correct Go | 0.175 (0.062) | 0.015 (0.026) | 0.746 (0.048) | 0.936 (0.053) |
| MID Large Rew vs Neu anticipation | 0.172 (0.055) | 0.012 (0.028) | 0.747 (0.04) | 0.939 (0.044) |
| SST Incorrect Stop vs Correct Go | 0.17 (0.085) | 0.015 (0.032) | 0.746 (0.051) | 0.936 (0.059) |
| T2 subcortical avg intensity | 0.157 (0.085) | 0.011 (0.033) | 0.755 (0.039) | 0.952 (0.043) |
| MID Small Rew vs Neu anticipation | 0.154 (0.086) | 0.007 (0.04) | 0.75 (0.04) | 0.941 (0.044) |
| MID Loss vs Neutral anticipation | 0.147 (0.07) | 0.004 (0.024) | 0.75 (0.04) | 0.942 (0.043) |
| SST Correct Go vs Fixation | 0.138 (0.065) | 0.005 (0.026) | 0.749 (0.046) | 0.938 (0.054) |
| SST Incorrect Go vs Correct Go | 0.122 (0.072) | 0.001 (0.03) | 0.752 (0.053) | 0.944 (0.059) |
| MID Large Loss vs Neu anticipation | 0.121 (0.074) | -0.004 (0.03) | 0.752 (0.04) | 0.942 (0.044) |
| T2 summations | 0.116 (0.07) | -0.003 (0.029) | 0.763 (0.041) | 0.96 (0.048) |
| MID Small Loss vs Neu Anticipation | 0.106 (0.071) | -0.005 (0.021) | 0.755 (0.041) | 0.948 (0.044) |
| SST Correct Stop vs Incorrect Stop | 0.09 (0.086) | -0.006 (0.023) | 0.754 (0.049) | 0.947 (0.057) |
| MID Large vs Small Loss Anticipation | 0.064 (0.07) | -0.012 (0.025) | 0.756 (0.043) | 0.948 (0.048) |
| MID Large vs Small Rew anticipation | 0.063 (0.059) | -0.012 (0.018) | 0.759 (0.042) | 0.952 (0.046) |
| SST Incorrect Go vs Incorrect Stop | 0.038 (0.067) | -0.014 (0.019) | 0.756 (0.052) | 0.95 (0.059) |
| ENback Positive vs Neutral Face | 0.006 (0.069) | -0.013 (0.018) | 0.732 (0.037) | 0.919 (0.044) |
| ENback Negative vs Neutral Face | -0.012 (0.031) | -0.012 (0.015) | 0.735 (0.039) | 0.923 (0.043) |
| ENback Emotion vs Neutral Face | -0.027 (0.067) | -0.014 (0.016) | 0.733 (0.038) | 0.921 (0.045) |

**Supplementary Table 4. Results of linear-mixed models using proxy measures of cognitive abilities based on mental health and/or neuroimaging as regressors to explain cognitive abilities across test sites in the baseline.** cws = values centred within each site; savg = values averaged within each site.

| Response | Cognitive abilities | | | Cognitive abilities | | | Cognitive abilities | | |
|---|---|---|---|---|---|---|---|---|---|
| *Regressors* | Estimates | CI | p | Estimates | CI | p | Estimates | CI | p |
| (Intercept) | 0.02 | -0.00 – 0.03 | 0.058 | 0.02 | -0.00 – 0.04 | 0.057 | 0.02 | -0.00 – 0.03 | 0.067 |
| mental savg | 0.00 | -0.02 – 0.02 | 0.895 | 0.00 | -0.02 – 0.02 | 0.985 | | | |
| mental cws | 0.19 | 0.17 – 0.20 | **<0.001** | 0.31 | 0.29 – 0.33 | **<0.001** | | | |
| neuroimaging savg | -0.01 | -0.02 – 0.01 | 0.507 | | | | -0.01 | -0.02 – 0.01 | 0.523 |
| neuroimaging cws | 0.43 | 0.41 – 0.44 | **<0.001** | | | | 0.48 | 0.47 – 0.50 | **<0.001** |
| Random Effects | | | | | | | | | |
| $\sigma^2$ | 0.55 | | | 0.54 | | | 0.57 | | |
| $\tau_{00}$ | 0.17 SITE_ID_L:REL_FAMILY_ID | | | 0.35 SITE_ID_L:REL_FAMILY_ID | | | 0.18 SITE_ID_L:REL_FAMILY_ID | | |
| ICC | 0.24 | | | 0.39 | | | 0.24 | | |
| N | 21 SITE_ID_L | | | 21 SITE_ID_L | | | 21 SITE_ID_L | | |
| | 9001 REL_FAMILY_ID | | | 9001 REL_FAMILY_ID | | | 9001 REL_FAMILY_ID | | |
| Observations | 10728 | | | 10728 | | | 10728 | | |
| Marginal $R^2$ | 0.272 | | | 0.098 | | | 0.238 | | |
| Conditional $R^2$ | 0.444 | | | 0.452 | | | 0.423 | | |

**Supplementary Table 5. Results of linear-mixed models using proxy measures of cognitive abilities based on mental health and/or neuroimaging as regressors to explain cognitive abilities across test sites in the follow-up.** cws = values centred within each site; savg = values averaged within each site.

| **Response** | **Cognitive abilities** | | | **Cognitive abilities** | | | **Cognitive abilities** | | |
|---|---|---|---|---|---|---|---|---|---|
| *Regressors* | Estimates | CI | p | Estimates | CI | p | Estimates | CI | p |
| (Intercept) | 0.82 | 0.80 – 0.84 | **<0.001** | 0.82 | 0.80 – 0.85 | **<0.001** | 0.82 | 0.80 – 0.84 | **<0.001** |
| mental savg | 0.02 | 0.00 – 0.04 | **0.047** | 0.02 | 0.00 – 0.05 | **0.037** | | | |
| mental cws | 0.19 | 0.17 – 0.21 | **<0.001** | 0.31 | 0.29 – 0.33 | **<0.001** | | | |
| neuroimaging savg | 0.02 | 0.00 – 0.05 | **0.021** | | | | 0.03 | 0.01 – 0.05 | **0.012** |
| neuroimaging cws | 0.42 | 0.40 – 0.44 | **<0.001** | | | | 0.47 | 0.45 – 0.49 | **<0.001** |
| Random Effects | | | | | | | | | |
| $\sigma^2$ | 0.41 | | | 0.45 | | | 0.42 | | |
| $\tau_{00}$ | 0.24 SITE_ID_L:REL_FAMILY_ID | | | 0.37 SITE_ID_L:REL_FAMILY_ID | | | 0.27 SITE_ID_L:REL_FAMILY_ID | | |
| ICC | 0.37 | | | 0.46 | | | 0.40 | | |
| N | 21 SITE_ID_L | | | 21 SITE_ID_L | | | 21 SITE_ID_L | | |
| | 5434 REL_FAMILY_ID | | | 5434 REL_FAMILY_ID | | | 5434 REL_FAMILY_ID | | |
| Observations | 6315 | | | 6315 | | | 6315 | | |
| Marginal $R^2$ | 0.286 | | | 0.104 | | | 0.245 | | |
| Conditional $R^2$ | 0.552 | | | 0.513 | | | 0.545 | | |

**Supplementary Table 6. Results of linear-mixed models using proxy measures of cognitive abilities based on mental health and/or polygenic scores as regressors to explain cognitive abilities across test sites in the baseline.** cws = values centred within each site; savg = values averaged within each site; cws,cwf = values centred within each family first and then within each site; savg,favg = values averaged within each family first and then within each site. PGS = polygenic scores.

| **Response** | **Cognitive abilities** | | | **Cognitive abilities** | | | **Cognitive abilities** | | |
|---|---|---|---|---|---|---|---|---|---|
| *Regressors* | Estimates | CI | p | Estimates | CI | p | Estimates | CI | p |
| (Intercept) | 0.23 | 0.21 – 0.26 | **<0.001** | 0.23 | 0.21 – 0.25 | **<0.001** | 0.23 | 0.21 – 0.26 | **<0.001** |
| mental savg | 0.06 | 0.02 – 0.09 | **0.004** | 0.13 | 0.10 – 0.15 | **<0.001** | | | |
| mental cws | 0.25 | 0.23 – 0.27 | **<0.001** | 0.25 | 0.23 – 0.27 | **<0.001** | | | |
| PGS savg favg | -0.08 | -0.12– -0.05 | **<0.001** | | | | -0.13 | -0.15 – -0.10 | **<0.001** |
| PGS cws cwf | 0.05 | 0.03 – 0.07 | **<0.001** | | | | 0.06 | 0.04 – 0.08 | **<0.001** |
| Random Effects | | | | | | | | | |
| $\sigma^2$ | 0.51 | | | 0.52 | | | 0.53 | | |
| $\tau_{00}$ | 0.27 $_{\text{SITE\_ID\_L:REL\_FAMILY\_ID}}$ | | | 0.26 $_{\text{SITE\_ID\_L:REL\_FAMILY\_ID}}$ | | | 0.32 $_{\text{SITE\_ID\_L:REL\_FAMILY\_ID}}$ | | |
| ICC | 0.34 | | | 0.33 | | | 0.38 | | |
| N | 21 $_{\text{SITE\_ID\_L}}$ | | | 21 $_{\text{SITE\_ID\_L}}$ | | | 21 $_{\text{SITE\_ID\_L}}$ | | |
| | 4734 $_{\text{REL\_FAMILY\_ID}}$ | | | 4734 $_{\text{REL\_FAMILY\_ID}}$ | | | 4734 $_{\text{REL\_FAMILY\_ID}}$ | | |
| Observations | 5766 | | | 5766 | | | 5766 | | |
| Marginal $R^2$ | 0.098 | | | 0.092 | | | 0.026 | | |
| Conditional $R^2$ | 0.408 | | | 0.394 | | | 0.394 | | |

**Supplementary Table 7. Results of linear-mixed models using proxy measures of cognitive abilities based on mental health and/or polygenic scores as regressors to explain cognitive abilities across test sites in the follow-up.** cws = values centred within each site; savg = values averaged within each site; cws,cwf = values centred within each family first and then within each site; savg,favg = values averaged within each family first and then within each site. PGS = polygenic scores.

| **Response** | **Cognitive abilities** | | | **Cognitive abilities** | | | **Cognitive abilities** | | |
|---|---|---|---|---|---|---|---|---|---|
| Predictors | Estimates | CI | p | Estimates | CI | p | Estimates | CI | p |
| (Intercept) | 1.06 | 1.03 – 1.09 | **<0.001** | 1.06 | 1.03 – 1.09 | **<0.001** | 1.06 | 1.03 – 1.09 | **<0.001** |
| mental savg | 0.03 | -0.00 – 0.07 | 0.063 | 0.07 | 0.05 – 0.10 | **<0.001** | | | |
| mental cws | 0.22 | 0.19 – 0.25 | **<0.001** | 0.22 | 0.20 – 0.25 | **<0.001** | | | |
| PGS savg favg | -0.07 | -0.10– -0.04 | **<0.001** | | | | -0.09 | -0.12 – -0.06 | **<0.001** |
| PGS cws cwf | 0.04 | 0.02 – 0.06 | **<0.001** | | | | 0.05 | 0.03 – 0.07 | **<0.001** |
| Random Effects | | | | | | | | | |
| $\sigma^2$ | 0.42 | | | 0.43 | | | 0.43 | | |
| $\tau_{00}$ | 0.32 SITE_ID_L:REL_FAMILY_ID | | | 0.31 SITE_ID_L:REL_FAMILY_ID | | | 0.37 SITE_ID_L:REL_FAMILY_ID | | |
| ICC | 0.43 | | | 0.42 | | | 0.46 | | |
| N | 21 SITE_ID_L | | | 21 SITE_ID_L | | | 21 SITE_ID_L | | |
| | 3370 REL_FAMILY_ID | | | 3370 REL_FAMILY_ID | | | 3370 REL_FAMILY_ID | | |
| Observations | 4036 | | | 4036 | | | 4036 | | |
| Marginal $R^2$ | 0.075 | | | 0.068 | | | 0.013 | | |
| Conditional $R^2$ | 0.470 | | | 0.460 | | | 0.469 | | |

**Supplementary Table 8. Results of linear-mixed models using proxy measures of cognitive abilities based on mental health and/or socio-demographics, lifestyles and developmental adverse events as regressors to explain cognitive abilities across test sites in the baseline.** cws = values centred within each site; savg = values averaged within each site; sdl= socio-demographics, lifestyles and developmental adverse events.

| **Response** | **Cognitive abilities** | | | **Cognitive abilities** | | | **Cognitive abilities** | | |
|---|---|---|---|---|---|---|---|---|---|
| *Regressors* | *Estimates* | *CI* | *p* | *Estimates* | *CI* | *p* | *Estimates* | *CI* | *p* |
| (Intercept) | 0.01 | -0.01 – 0.02 | 0.525 | 0.01 | -0.01 – 0.03 | 0.385 | 0.01 | -0.01 – 0.02 | 0.558 |
| mental savg | -0.00 | -0.02 – 0.02 | 0.917 | -0.00 | -0.02 – 0.02 | 0.930 | | | |
| mental cws | 0.20 | 0.18 – 0.22 | **<0.001** | 0.31 | 0.29 – 0.33 | **<0.001** | | | |
| sdl savg | 0.00 | -0.02 – 0.02 | 0.819 | | | | 0.00 | -0.01 – 0.02 | 0.792 |
| sdl cws | 0.40 | 0.38 – 0.41 | **<0.001** | | | | 0.46 | 0.44 – 0.48 | **<0.001** |
| **Random Effects** | | | | | | | | | |
| $\sigma^2$ | 0.52 | | | 0.53 | | | 0.54 | | |
| $\tau_{00}$ | 0.22 SITE_ID_L:REL_FAMILY_ID | | | 0.35 SITE_ID_L:REL_FAMILY_ID | | | 0.24 SITE_ID_L:REL_FAMILY_ID | | |
| ICC | 0.30 | | | 0.40 | | | 0.31 | | |
| N | 21 SITE_ID_L | | | 21 SITE_ID_L | | | 21 SITE_ID_L | | |
| | 9390 REL_FAMILY_ID | | | 9390 REL_FAMILY_ID | | | 9390 REL_FAMILY_ID | | |
| Observations | 11294 | | | 11294 | | | 11294 | | |
| Marginal $R^2$ | 0.249 | | | 0.098 | | | 0.213 | | |
| Conditional $R^2$ | 0.474 | | | 0.458 | | | 0.456 | | |

**Supplementary Table 9. Results of linear-mixed models using proxy measures of cognitive abilities based on mental health and/or socio-demographics, lifestyles and developmental adverse events as regressors to explain cognitive abilities across test sites in the follow-up.** cws = values centred within each site; savg = values averaged within each site; sdl= socio-demographics, lifestyles and developmental adverse events.

| **Response** | **Cognitive abilities** | | | **Cognitive abilities** | | | **Cognitive abilities** | | |
|---|---|---|---|---|---|---|---|---|---|
| *Regressors* | *Estimates* | *CI* | *p* | *Estimates* | *CI* | *p* | *Estimates* | *CI* | *p* |
| (Intercept) | 0.83 | 0.81 –0.85 | **<0.001** | 0.83 | 0.81 –0.86 | **<0.001** | 0.83 | 0.81 –0.85 | **<0.001** |
| mental savg | 0.01 | -0.01–0.03 | 0.185 | 0.01 | -0.01–0.04 | 0.198 | | | |
| mental cws | 0.20 | 0.18 – 0.22 | **<0.001** | 0.30 | 0.28 – 0.32 | **<0.001** | | | |
| sdl savg | 0.00 | -0.02–0.02 | 0.957 | | | | 0.00 | -0.02 – 0.02 | 0.757 |
| sdl cws | 0.39 | 0.37 – 0.41 | **<0.001** | | | | 0.44 | 0.42 – 0.47 | **<0.001** |
| **Random Effects** | | | | | | | | | |
| $\sigma^2$ | 0.42 | | | 0.45 | | | 0.43 | | |
| $\tau_{00}$ | 0.27 SITE_ID_L:REL_FAMILY_ID | | | 0.37 SITE_ID_L:REL_FAMILY_ID | | | 0.30 SITE_ID_L:REL_FAMILY_ID | | |
| ICC | 0.39 | | | 0.45 | | | 0.41 | | |
| N | 21 SITE_ID_L | | | 21 SITE_ID_L | | | 21 SITE_ID_L | | |
| | 6217 REL_FAMILY_ID | | | 6217 REL_FAMILY_ID | | | 6217 REL_FAMILY_ID | | |
| Observations | 7382 | | | 7382 | | | 7382 | | |
| Marginal $R^2$ | 0.256 | | | 0.099 | | | 0.213 | | |
| Conditional $R^2$ | 0.543 | | | 0.508 | | | 0.535 | | |

**Supplementary Table 10. Results of linear-mixed models using proxy measures of cognitive abilities based on mental health, neuroimaging, polygenic scores and/or socio-demographics, lifestyles and developmental adverse events as regressors to explain cognitive abilities across test sites in the baseline.** cws = values centred within each site; savg = values averaged within each site; cws,cwf = values centred within each family first and then within each site; savg,favg = values averaged within each family first and then within each site; PGS = polygenic scores; sdl= socio-demographics, lifestyles and developmental adverse events.

| **Response** | **Cognitive abilities** | | | **Cognitive abilities** | | |
|---|---|---|---|---|---|---|
| *Regressors* | Estimates | CI | p | Estimates | CI | p |
| (Intercept) | 0.24 | 0.21 – 0.26 | **<0.001** | 0.24 | 0.21 – 0.26 | **<0.001** |
| mental savg | 0.00 | -0.05 – 0.05 | 0.975 | 0.09 | 0.05 – 0.12 | **<0.001** |
| mental cws | 0.14 | 0.11 – 0.16 | **<0.001** | 0.18 | 0.15 – 0.20 | **<0.001** |
| neuroimaging savg | 0.01 | -0.03 – 0.05 | 0.533 | 0.05 | 0.01 – 0.09 | **0.006** |
| neuroimaging cws | 0.26 | 0.24 – 0.29 | **<0.001** | 0.31 | 0.28 – 0.33 | **<0.001** |
| PGS savg favg | -0.04 | -0.08 – 0.00 | 0.070 | | | |
| PGS cws cwf | 0.05 | 0.03 – 0.07 | **<0.001** | | | |
| sdl savg | 0.09 | 0.03 – 0.16 | **0.006** | | | |
| sdl cws | 0.18 | 0.16 – 0.21 | **<0.001** | | | |
| $\sigma^2$ | 0.50 | | | 0.52 | | |
| $\tau_{00}$ | 0.15 SITE_ID_L:REL_FAMILY_ID | | | 0.17 SITE_ID_L:REL_FAMILY_ID | | |
| ICC | 0.23 | | | 0.25 | | |
| N | 21 SITE_ID_L | | | 21 SITE_ID_L | | |
| | 4571 REL_FAMILY_ID | | | 4571 REL_FAMILY_ID | | |
| Observations | 5520 | | | 5520 | | |
| Marginal $R^2$ | 0.241 | | | 0.197 | | |
| Conditional $R^2$ | 0.416 | | | 0.395 | | |

| *Regressors* | Estimates | CI | p | Estimates | CI | p |
|---|---|---|---|---|---|---|
| (Intercept) | 0.24 | 0.21 – 0.26 | **<0.001** | 0.24 | 0.21 – 0.26 | <0.001 |
| mental savg | 0.06 | 0.03 – 0.10 | **0.001** | 0.00 | -0.04 – 0.05 | 0.890 |
| mental cws | 0.24 | 0.22 – 0.27 | **<0.001** | 0.19 | 0.16 – 0.21 | <0.001 |
| neuroimaging savg | | | | | | |
| neuroimaging cws | | | | | | |
| PGS savg favg | -0.08 | -0.12–-0.05 | **<0.001** | | | |
| PGS cws cwf | 0.06 | 0.04 – 0.08 | **<0.001** | | | |
| sdl savg | | | | 0.14 | 0.09 – 0.19 | <0.001 |
| sdl cws | | | | 0.25 | 0.22 – 0.27 | <0.001 |
| $\sigma^2$ | 0.51 | | | 0.52 | | |
| $\tau_{00}$ | 0.27 SITE_ID_L:REL_FAMILY_ID | | | 0.20 SITE_ID_L:REL_FAMILY_ID | | |
| ICC | 0.34 | | | 0.28 | | |
| N | 21 SITE_ID_L | | | 21 SITE_ID_L | | |
| | 4571 REL_FAMILY_ID | | | 4571 REL_FAMILY_ID | | |
| Observations | 5520 | | | 5520 | | |
| Marginal $R^2$ | 0.097 | | | 0.163 | | |
| Conditional $R^2$ | 0.408 | | | 0.395 | | |

**Supplementary Table 11. Results of linear-mixed models using proxy measures of cognitive abilities based on mental health, neuroimaging, polygenic scores and/or socio-demographics, lifestyles and developmental adverse events as regressors to explain cognitive abilities across test sites in the follow-up.** cws = values centred within each site; savg = values averaged within each site; cws,cwf = values centred within each family first and then within each site; savg,favg = values averaged within each family first and then within each site; PGS = polygenic scores; sdl= socio-demographics, lifestyles and developmental adverse events.

| **Response** | **Cognitive abilities** | | | **Cognitive abilities** | | |
|---|---|---|---|---|---|---|
| *Regressors* | *Estimates* | *CI* | *p* | *Estimates* | *CI* | *p* |
| (Intercept) | 1.05 | 1.02 – 1.08 | **<0.001** | 1.05 | 1.02 – 1.08 | **<0.001** |
| mental savg | 0.05 | -0.01 – 0.10 | 0.100 | 0.06 | 0.03 – 0.10 | **<0.001** |
| mental cws | 0.13 | 0.11 – 0.16 | **<0.001** | 0.17 | 0.14 – 0.20 | **<0.001** |
| neuroimaging savg | 0.00 | -0.06 – 0.06 | 0.935 | 0.03 | -0.01 – 0.06 | 0.146 |
| neuroimaging cws | 0.27 | 0.24 – 0.30 | **<0.001** | 0.31 | 0.28 – 0.33 | **<0.001** |
| PGS savg favg | 0.00 | -0.03 – 0.04 | 0.833 | | | |
| PGS cws cwf | 0.04 | 0.02 – 0.06 | **<0.001** | | | |
| sdl savg | 0.04 | -0.04 – 0.12 | 0.349 | | | |
| sdl cws | 0.20 | 0.17 – 0.23 | **<0.001** | | | |
| $\sigma^2$ | 0.38 | | | 0.40 | | |
| $\tau_{00}$ | 0.23 SITE_ID_L:REL_FAMILY_ID | | | 0.25 SITE_ID_L:REL_FAMILY_ID | | |
| ICC | 0.38 | | | 0.39 | | |
| N | 21 SITE_ID_L | | | 21 SITE_ID_L | | |
| | 2930 REL_FAMILY_ID | | | 2930 REL_FAMILY_ID | | |
| Observations | 3423 | | | 3423 | | |
| Marginal $R^2$ / Conditional $R^2$ | 0.242 / 0.527 | | | 0.190 / 0.506 | | |

| *Regressors* | *Estimates* | *CI* | *p* | *Estimates* | *CI* | *p* |
|---|---|---|---|---|---|---|
| (Intercept) | 1.05 | 1.02 – 1.08 | **<0.001** | 1.05 | 1.02 – 1.08 | **<0.001** |
| mental savg | 0.08 | 0.04 – 0.11 | **<0.001** | 0.05 | -0.00 – 0.10 | 0.074 |
| mental cws | 0.23 | 0.20 – 0.26 | **<0.001** | 0.18 | 0.15 – 0.21 | **<0.001** |
| neuroimaging savg | | | | | | |
| neuroimaging cws | | | | | | |
| PGS savg favg | 0.00 | - 0.03 – 0.04 | 0.844 | | | |
| PGS cws cwf | 0.05 | 0.03 – 0.07 | **<0.001** | | | |
| sdl savg | | | | 0.04 | -0.01 – 0.09 | 0.092 |
| sdl cws | | | | 0.25 | 0.22 – 0.28 | **<0.001** |
| $\sigma^2$ | 0.41 | | | 0.42 | | |
| $\tau_{00}$ | 0.33 SITE_ID_L:REL_FAMILY_ID | | | 0.27 SITE_ID_L:REL_FAMILY_ID | | |
| ICC | 0.45 | | | 0.39 | | |
| N | 21 SITE_ID_L | | | 21 SITE_ID_L | | |
| | 2930 REL_FAMILY_ID | | | 2930 REL_FAMILY_ID | | |
| Observations | 3423 | | | 3423 | | |
| Marginal $R^2$ / Conditional $R^2$ | 0.076 / 0.491 | | | 0.153 / 0.486 | | |

**Supplementary Table 12. Exclusion criteria for neuroimaging features in the baseline.**

| Neuroimaging features | Data provided | Did not pass quality control | Had vision problems | From site 22 | Had any missing feature | Flagged as outliers | Observations kept |
|---|---|---|---|---|---|---|---|
| ENback 0back | 11771 | 3996 | 38 | 21 | 8 | 292 | 7416 |
| ENback 2back | 11771 | 3996 | 38 | 21 | 12 | 281 | 7423 |
| ENback 2back vs 0back | 11771 | 3996 | 38 | 21 | 10 | 397 | 7309 |
| ENback emotion | 11771 | 3996 | 38 | 21 | 10 | 303 | 7403 |
| ENback Emotion vs Neutral Face | 11771 | 3996 | 38 | 21 | 11 | 480 | 7225 |
| ENback Face vs Place | 11771 | 3996 | 38 | 21 | 10 | 391 | 7315 |
| ENback Negative vs Neutral Face | 11771 | 3996 | 38 | 21 | 11 | 454 | 7251 |
| ENback Positive vs Neutral Face | 11771 | 3996 | 38 | 21 | 10 | 500 | 7206 |
| ENback place | 11771 | 3996 | 38 | 21 | 11 | 331 | 7374 |
| MID Reward vs Neutral anticipation | 11771 | 2596 | 51 | 22 | 11 | 250 | 8841 |
| MID Loss vs Neutral anticipation | 11771 | 2596 | 51 | 22 | 11 | 245 | 8846 |
| MID Positive vs Negative Reward Feedback | 11771 | 2596 | 51 | 22 | 12 | 338 | 8752 |
| MID Positive vs Negative Punishment Feedback | 11771 | 2596 | 51 | 22 | 10 | 334 | 8758 |
| MID Large Reward vs Neutral anticipation | 11771 | 2596 | 51 | 22 | 12 | 241 | 8849 |
| MID Small Reward vs Neutral anticipation | 11771 | 2596 | 51 | 22 | 10 | 270 | 8822 |
| MID Large Reward vs Small Reward anticipation | 11771 | 2596 | 51 | 22 | 13 | 266 | 8823 |
| MID Large Loss vs Neutral anticipation | 11771 | 2596 | 51 | 22 | 11 | 250 | 8841 |
| MID Smal Loss vs Neutral anticipation | 11771 | 2596 | 51 | 22 | 11 | 282 | 8809 |
| MID Large Loss vs Small Loss anticipation | 11771 | 2596 | 51 | 22 | 12 | 307 | 8783 |
| SST Any Stop vs Correct Go | 11771 | 3672 | 45 | 20 | 14 | 227 | 7793 |
| SST Correct Go vs Fixation | 11771 | 3672 | 45 | 20 | 13 | 262 | 7759 |
| SST Correct Stop vs Correct Go | 11771 | 3672 | 45 | 20 | 13 | 236 | 7785 |
| SST Correct Stop vs Incorrect Stop | 11771 | 3672 | 45 | 20 | 14 | 292 | 7728 |
| SST Incorrect Go vs Correct Go | 11771 | 3672 | 45 | 20 | 15 | 481 | 7538 |
| SST Incorrect Go vs Incorrect Stop | 11771 | 3672 | 45 | 20 | 14 | 366 | 7654 |
| SST Incorrect Stop vs Correct Go | 11771 | 3672 | 45 | 20 | 13 | 246 | 7775 |
| rsfMRI temporal variance | 11771 | 2397 | 62 | 25 | 14 | 682 | 8591 |
| rsfMRI subcortical-network FC | 11771 | 2397 | 62 | 25 | 14 | 1 | 9272 |
| rsfMRI cortical FC | 11771 | 2397 | 62 | 25 | 14 | 3 | 9270 |
| T1 subcortical avg intensity | 11771 | 501 | 66 | 27 | 0 | 60 | 11117 |
| T1 white matter avg intensity | 11771 | 501 | 66 | 27 | 12 | 13 | 11152 |
| T1 gray matter avg intensity | 11771 | 501 | 66 | 27 | 12 | 11 | 11154 |
| T1 normalised intensity | 11771 | 501 | 66 | 27 | 12 | 2 | 11163 |
| T1 summations | 11771 | 501 | 66 | 27 | 12 | 34 | 11131 |
| cortical thickness | 11771 | 501 | 66 | 27 | 12 | 2 | 11163 |
| cortical area | 11771 | 501 | 66 | 27 | 12 | 1 | 11164 |
| cortical volume | 11771 | 501 | 66 | 27 | 12 | 0 | 11165 |
| subcortical volume | 11771 | 501 | 66 | 27 | 0 | 215 | 10962 |
| sulcal depth | 11771 | 501 | 66 | 27 | 12 | 1106 | 10059 |
| T2 subcortical avg intensity | 11771 | 1217 | 58 | 25 | 0 | 67 | 10404 |
| T2 white matter avg intensity | 11771 | 1217 | 58 | 25 | 10 | 56 | 10405 |
| T2 gray matter avg intensity | 11771 | 1217 | 58 | 25 | 10 | 55 | 10406 |
| T2 normalised intensity | 11771 | 1217 | 58 | 25 | 10 | 12 | 10449 |
| T2 summations | 11771 | 1217 | 58 | 25 | 10 | 14 | 10447 |
| DTI | 11771 | 1577 | 57 | 13 | 0 | 24 | 10100 |

**Supplementary Table 13. Exclusion criteria for neuroimaging features in the follow up.**

| Neuroimaging Features | Data provided | Did not pass quality control | Had vision problems | Had any missing feature | Flagged as outliers | Observations kept |
|---|---|---|---|---|---|---|
| ENback 0back | 8123 | 1804 | 35 | 11 | 216 | 6057 |
| ENback 2back | 8123 | 1804 | 35 | 13 | 186 | 6085 |
| ENback 2back vs 0back | 8123 | 1804 | 35 | 14 | 294 | 5976 |
| ENback emotion | 8123 | 1804 | 35 | 13 | 202 | 6069 |
| ENback Emotion vs Neutral Face | 8123 | 1804 | 35 | 13 | 347 | 5924 |
| ENback Face vs Place | 8123 | 1804 | 35 | 13 | 295 | 5976 |
| ENback Negative vs Neutral Face | 8123 | 1804 | 35 | 11 | 355 | 5918 |
| ENback Positive vs Neutral Face | 8123 | 1804 | 35 | 13 | 342 | 5929 |
| ENback place | 8123 | 1804 | 35 | 12 | 234 | 6038 |
| MID Reward vs Neutral anticipation | 8123 | 1379 | 40 | 8 | 153 | 6543 |
| MID Loss vs Neutral anticipation | 8123 | 1379 | 40 | 8 | 154 | 6542 |
| MID Positive vs Negative Reward Feedback | 8123 | 1379 | 40 | 9 | 192 | 6503 |
| MID Positive vs Negative Punishment Feedback | 8123 | 1379 | 40 | 9 | 197 | 6498 |
| MID Large Reward vs Neutral anticipation | 8123 | 1379 | 40 | 8 | 142 | 6554 |
| MID Small Reward vs Neutral anticipation | 8123 | 1379 | 40 | 8 | 163 | 6533 |
| MID Large Reward vs Small Reward anticipation | 8123 | 1379 | 40 | 8 | 155 | 6541 |
| MID Large Loss vs Neutral anticipation | 8123 | 1379 | 40 | 8 | 150 | 6546 |
| MID Smal Loss vs Neutral anticipation | 8123 | 1379 | 40 | 9 | 179 | 6516 |
| MID Large Loss vs Small Loss anticipation | 8123 | 1379 | 40 | 9 | 173 | 6522 |
| SST Any Stop vs Correct Go | 8123 | 2036 | 33 | 7 | 123 | 5924 |
| SST Correct Go vs Fixation | 8123 | 2036 | 33 | 7 | 173 | 5874 |
| SST Correct Stop vs Correct Go | 8123 | 2036 | 33 | 7 | 163 | 5884 |
| SST Correct Stop vs Incorrect Stop | 8123 | 2036 | 33 | 7 | 187 | 5860 |
| SST Incorrect Go vs Correct Go | 8123 | 2036 | 33 | 7 | 345 | 5702 |
| SST Incorrect Go vs Incorrect Stop | 8123 | 2036 | 33 | 7 | 267 | 5780 |
| SST Incorrect Stop vs Correct Go | 8123 | 2036 | 33 | 7 | 131 | 5916 |
| rsfMRI temporal variance | 8123 | 1152 | 49 | 14 | 512 | 6396 |
| rsfMRI subcortical-network FC | 8123 | 1152 | 49 | 14 | 3 | 6905 |
| rsfMRI cortical FC | 8123 | 1152 | 49 | 14 | 3 | 6905 |
| T1 subcortical avg intensity | 8123 | 227 | 51 | 0 | 32 | 7813 |
| T1 white matter avg intensity | 8123 | 227 | 51 | 10 | 8 | 7827 |
| T1 gray matter avg intensity | 8123 | 227 | 51 | 10 | 9 | 7826 |
| T1 normalised intensity | 8123 | 227 | 51 | 10 | 0 | 7835 |
| T1 summations | 8123 | 227 | 51 | 10 | 19 | 7816 |
| cortical thickness | 8123 | 227 | 51 | 10 | 2 | 7833 |
| cortical area | 8123 | 227 | 51 | 10 | 0 | 7835 |
| cortical volume | 8123 | 227 | 51 | 10 | 0 | 7835 |
| subcortical volume | 8123 | 227 | 51 | 0 | 112 | 7733 |
| sulcal depth | 8123 | 227 | 51 | 10 | 890 | 6945 |
| T2 subcortical avg intensity | 8123 | 600 | 50 | 0 | 39 | 7434 |
| T2 white matter avg intensity | 8123 | 600 | 50 | 10 | 47 | 7416 |
| T2 gray matter avg intensity | 8123 | 600 | 50 | 10 | 49 | 7414 |
| T2 normalised intensity | 8123 | 600 | 50 | 10 | 5 | 7458 |
| T2 summations | 8123 | 600 | 50 | 10 | 14 | 7449 |
| DTI | 8123 | 638 | 47 | 0 | 15 | 7423 |

**Supplementary Table 14. Summary statistics of the measures of mental health in the baseline.** Med = Median IQR = interquartile range; CV = Coefficient of variation; CBCL = Child Behavioural Checklist, reflecting children's emotional and behavioural problems; UPPS-P = Urgency, Premeditation, Perseverance, Sensation seeking and Positive urgency Impulsive Behaviour Scale; BAS = Behavioural Activation System. Under the variable names, there are information about the method to compute these variables and the original variables names in ABCD data dictionary.

| No | Variable | Stats / Values | Graph | Valid |
|---|---|---|---|---|
| 1 | Anxious/Depressed CBCL<br>[numeric]<br>CBCL_SCR_SYN_ANXDEP_R | Mean (sd) : 2.5 (3.1)<br>min < med < max:<br>0 < 1 < 26<br>IQR (CV) : 4 (1.2) | 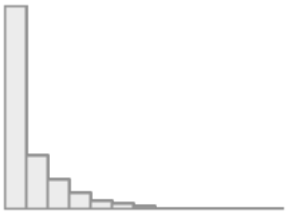 | 11754<br>(99.9%) |
| 2 | Withdrawn CBCL<br>[numeric]<br>CBCL_SCR_SYN_WITHDEP_R | Mean (sd) : 1 (1.7)<br>min < med < max:<br>0 < 0 < 15<br>IQR (CV) : 1 (1.7) | 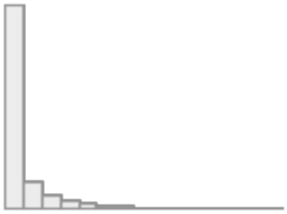 | 11754<br>(99.9%) |
| 3 | Somatic CBCL<br>[numeric]<br>CBCL_SCR_SYN_SOMATIC_R | Mean (sd) : 1.5 (2)<br>min < med < max:<br>0 < 1 < 16<br>IQR (CV) : 2 (1.3) | 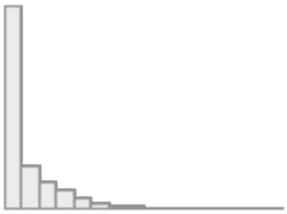 | 11754<br>(99.9%) |
| 4 | Social CBCL<br>[numeric]<br>CBCL_SCR_SYN_SOCIAL_R | Mean (sd) : 1.6 (2.3)<br>min < med < max:<br>0 < 1 < 18<br>IQR (CV) : 2 (1.4) | 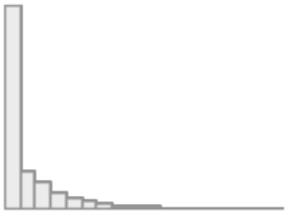 | 11754<br>(99.9%) |
| 5 | Thought CBCL<br>[numeric]<br>CBCL_SCR_SYN_THOUGHT_R | Mean (sd) : 1.6 (2.2)<br>min < med < max:<br>0 < 1 < 18<br>IQR (CV) : 2 (1.4) | 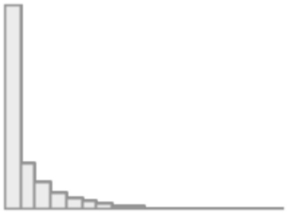 | 11754<br>(99.9%) |
| 6 | Attention CBCL<br>[numeric]<br>CBCL_SCR_SYN_ATTENTION_R | Mean (sd) : 3 (3.5)<br>min < med < max:<br>0 < 2 < 20<br>IQR (CV) : 5 (1.2) | 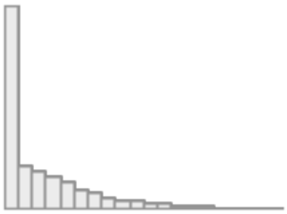 | 11754<br>(99.9%) |
| 7 | Rule-Breaking CBCL<br>[numeric]<br>CBCL_SCR_SYN_RULEBREAK_R | Mean (sd) : 1.2 (1.9)<br>min < med < max:<br>0 < 0 < 20<br>IQR (CV) : 2 (1.6) | 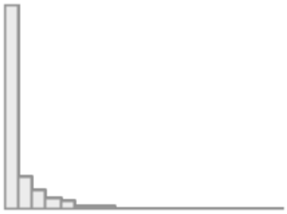 | 11754<br>(99.9%) |

| No | Variable | Stats / Values | Graph | Valid |
|---|---|---|---|---|
| 8 | Aggressive CBCL<br>[numeric]<br>CBCL_SCR_SYN_AGGRESSIVE_R | Mean (sd) : 3.3 (4.4)<br>min < med < max:<br>0 < 2 < 36<br>IQR (CV) : 5 (1.3) | 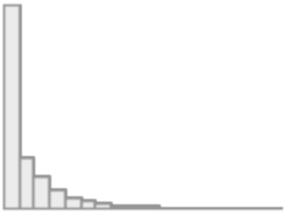 | 11754<br>(99.9%) |
| 9 | Negative Urgency UPSS<br>[numeric]<br>UPPS_Y_SS_NEGATIVE_URGENCY | Mean (sd) : 8.5 (2.6)<br>min < med < max:<br>4 < 8 < 16<br>IQR (CV) : 3 (0.3) | 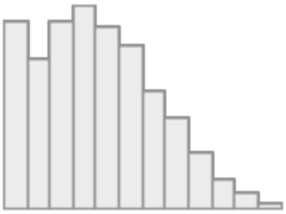 | 11739<br>(99.8%) |
| 10 | Lack of Planning UPSS<br>[numeric]<br>UPPS_Y_SS_LACK_OF_PLANNING | Mean (sd) : 7.7 (2.4)<br>min < med < max:<br>4 < 8 < 16<br>IQR (CV) : 3 (0.3) | 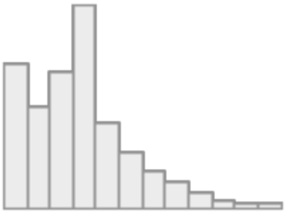 | 11739<br>(99.8%) |
| 11 | Sensation Seeking UPSS<br>[numeric]<br>UPPS_Y_SS_SENSATION_SEEKING | Mean (sd) : 9.8 (2.7)<br>min < med < max:<br>4 < 10 < 16<br>IQR (CV) : 4 (0.3) | 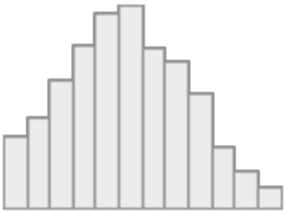 | 11739<br>(99.8%) |
| 12 | Positive Urgency UPSS<br>[numeric]<br>UPPS_Y_SS_POSITIVE_URGENCY | Mean (sd) : 8 (3)<br>min < med < max:<br>4 < 8 < 16<br>IQR (CV) : 4 (0.4) | 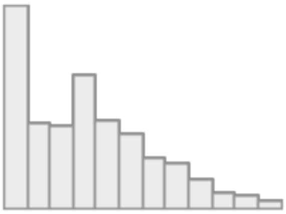 | 11739<br>(99.8%) |
| 13 | Lack of Perseverance UPSS<br>[numeric]<br>UPPS_Y_SS_LACK_OF_PERSEVERANCE | Mean (sd) : 7 (2.3)<br>min < med < max:<br>4 < 7 < 16<br>IQR (CV) : 3 (0.3) | 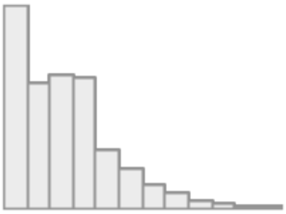 | 11739<br>(99.8%) |
| 14 | Behavioural Inhibition System<br>[numeric]<br>Mean of<br>(BISBAS2_Y,<br>BISBAS3_Y,<br>BISBAS4_Y,<br>BISBAS6_Y) | Mean (sd) : 1.4 (0.7)<br>min < med < max:<br>0 < 1.2 < 3<br>IQR (CV) : 1.2 (0.5) | 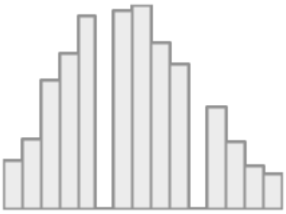 | 11740<br>(99.8%) |
| 15 | BAS Reward Responsiveness<br>[numeric]<br>Mean of<br>(BISBAS8_Y,<br>BISBAS10_Y,<br>BISBAS11_Y,<br>BISBAS12_Y) | Mean (sd) : 2.2 (0.6)<br>min < med < max:<br>0 < 2.2 < 3<br>IQR (CV) : 1 (0.3) | 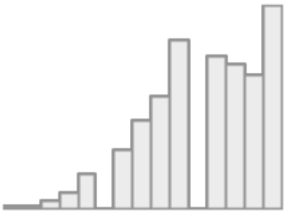 | 11739<br>(99.8%) |

| No | Variable | Stats / Values | Graph | Valid |
|---|---|---|---|---|
| 16 | BAS Drive<br>[numeric]<br>Mean of<br>(BISBAS13_Y,<br>BISBAS14_Y,<br>BISBAS15_Y,<br>BISBAS16_Y) | Mean (sd) : 1 (0.8)<br>min < med < max:<br>0 < 1 < 3<br>IQR (CV) : 1 (0.7) | 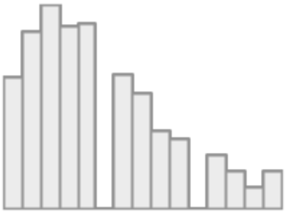 | 11739<br>(99.8%) |
| 17 | BAS Fun<br>[numeric]<br>Means of<br>(BISBAS17_Y,<br>BISBAS18_Y,<br>BISBAS19_Y,<br>BISBAS20_Y) | Mean (sd) : 1.4 (0.7)<br>min < med < max:<br>0 < 1.5 < 3<br>IQR (CV) : 0.8 (0.5) | 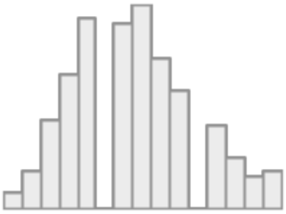 | 11739<br>(99.8%) |

**Supplementary Table 15. Summary statistics of the measures of mental health in the follow up.** Med = Median IQR = interquartile range; CV = Coefficient of variation; CBCL = Child Behavioural Checklist, reflecting children's emotional and behavioural problems; UPPS-P = Urgency, Premeditation, Perseverance, Sensation seeking and Positive urgency Impulsive Behaviour Scale; BAS = Behavioural Activation System. Under the variable names, there are information about the method to compute these variables and the original variables names in ABCD data dictionary.

| No | Variable | Stats / Values | Graph | Valid |
|---|---|---|---|---|
| 1 | Anxious/Depressed CBCL<br>[numeric]<br>CBCL_SCR_SYN_ANXDEP_R | Mean (sd) : 2.3 (3)<br>min < med < max:<br>0 < 1 < 24<br>IQR (CV) : 3 (1.3) | 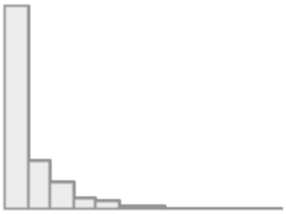 | 10827<br>(99.9%) |
| 2 | Withdrawn CBCL<br>[numeric]<br>CBCL_SCR_SYN_WITHDEP_R | Mean (sd) : 1.3 (2)<br>min < med < max:<br>0 < 0 < 16<br>IQR (CV) : 2 (1.6) | 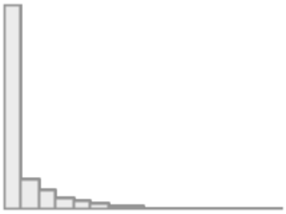 | 10827<br>(99.9%) |
| 3 | Somatic CBCL<br>[numeric]<br>CBCL_SCR_SYN_SOMATIC_R | Mean (sd) : 1.4 (1.9)<br>min < med < max:<br>0 < 1 < 16<br>IQR (CV) : 2 (1.4) | 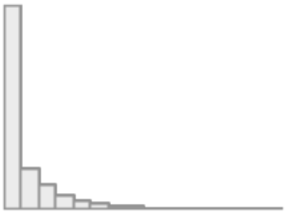 | 10827<br>(99.9%) |
| 4 | Social CBCL<br>[numeric]<br>CBCL_SCR_SYN_SOCIAL_R | Mean (sd) : 1.3 (2.1)<br>min < med < max:<br>0 < 0 < 17<br>IQR (CV) : 2 (1.6) | 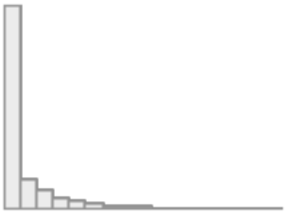 | 10827<br>(99.9%) |
| 5 | Thought CBCL<br>[numeric]<br>CBCL_SCR_SYN_THOUGHT_R | Mean (sd) : 1.4 (2.1)<br>min < med < max:<br>0 < 1 < 22<br>IQR (CV) : 2 (1.5) | 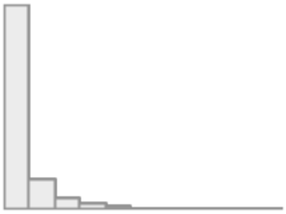 | 10827<br>(99.9%) |
| 6 | Attention CBCL<br>[numeric]<br>CBCL_SCR_SYN_ATTENTION_R | Mean (sd) : 2.7 (3.3)<br>min < med < max:<br>0 < 1 < 19<br>IQR (CV) : 4 (1.2) | 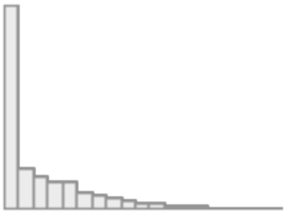 | 10827<br>(99.9%) |
| 7 | Rule-Breaking CBCL<br>[numeric]<br>CBCL_SCR_SYN_RULEBREAK_R | Mean (sd) : 1.1 (1.9)<br>min < med < max:<br>0 < 0 < 23<br>IQR (CV) : 1 (1.7) | 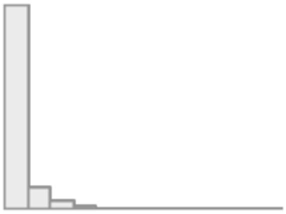 | 10827<br>(99.9%) |

| No | Variable | Stats / Values | Graph | Valid |
|---|---|---|---|---|
| 8 | Aggressive CBCL<br>[numeric]<br>CBCL_SCR_SYN_AGGRESSIVE_R | Mean (sd) : 2.8 (4)<br>min < med < max:<br>0 < 1 < 33<br>IQR (CV) : 4 (1.4) | 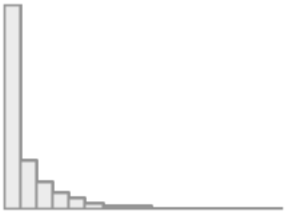 | 10827<br>(99.9%) |
| 9 | Negative Urgency UPSS<br>[numeric]<br>UPPS_Y_SS_NEGATIVE_URGENCY | Mean (sd) : 7.8 (2.3)<br>min < med < max:<br>4 < 8 < 16<br>IQR (CV) : 3 (0.3) | 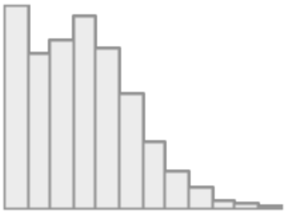 | 10811<br>(99.7%) |
| 10 | Lack of Planning UPSS<br>[numeric]<br>UPPS_Y_SS_LACK_OF_PLANNING | Mean (sd) : 7.8 (2.2)<br>min < med < max:<br>4 < 8 < 16<br>IQR (CV) : 3 (0.3) | 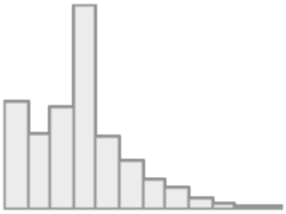 | 10811<br>(99.7%) |
| 11 | Sensation Seeking UPSS<br>[numeric]<br>UPPS_Y_SS_SENSATION_SEEKING | Mean (sd) : 9.5 (2.7)<br>min < med < max:<br>3 < 9 < 16<br>IQR (CV) : 3 (0.3) | 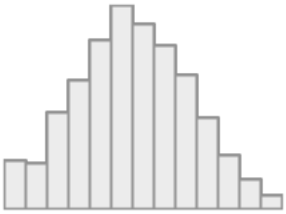 | 10812<br>(99.7%) |
| 12 | Positive Urgency UPSS<br>[numeric]<br>UPPS_Y_SS_POSITIVE_URGENCY | Mean (sd) : 7.4 (2.7)<br>min < med < max:<br>4 < 8 < 16<br>IQR (CV) : 4 (0.4) | 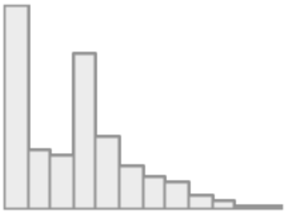 | 10811<br>(99.7%) |
| 13 | Lack of Perseverance UPSS<br>[numeric]<br>UPPS_Y_SS_LACK_OF_PERSEVERANCE | Mean (sd) : 7 (2.3)<br>min < med < max:<br>4 < 7 < 16<br>IQR (CV) : 3 (0.3) | 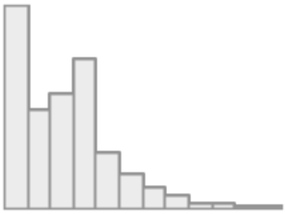 | 10811<br>(99.7%) |
| 14 | Behavioural Inhibition System<br>[numeric]<br>Mean of<br>(BISBAS2_Y,<br>BISBAS3_Y,<br>BISBAS4_Y,<br>BISBAS6_Y) | Mean (sd) : 1.2 (0.7)<br>min < med < max:<br>0 < 1.2 < 3<br>IQR (CV) : 1 (0.6) | 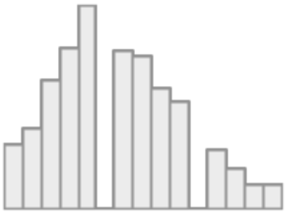 | 10811<br>(99.7%) |
| 15 | BAS Reward Responsiveness<br>[numeric]<br>Mean of<br>(BISBAS8_Y,<br>BISBAS10_Y,<br>BISBAS11_Y,<br>BISBAS12_Y) | Mean (sd) : 2 (0.6)<br>min < med < max:<br>0 < 2 < 3<br>IQR (CV) : 1 (0.3) | 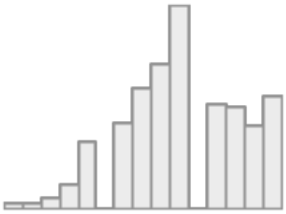 | 10811<br>(99.7%) |

| No | Variable | Stats / Values | Graph | Valid |
|---|---|---|---|---|
| 16 | BAS Drive<br>[numeric]<br>Mean of<br>(BISBAS13_Y,<br>BISBAS14_Y,<br>BISBAS15_Y,<br>BISBAS16_Y) | Mean (sd) : 0.9 (0.7)<br>min < med < max:<br>0 < 0.8 < 3<br>IQR (CV) : 0.8 (0.8) | 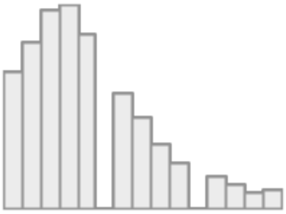 | 10811<br>(99.7%) |
| 17 | BAS Fun<br>[numeric]<br>Mean of<br>(BISBAS17_Y,<br>BISBAS18_Y,<br>BISBAS19_Y,<br>BISBAS20_Y) | Mean (sd) : 1.1 (0.6)<br>min < med < max:<br>0 < 1 < 3<br>IQR (CV) : 0.8 (0.6) | 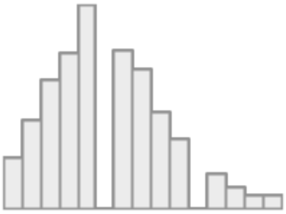 | 10811<br>(99.7%) |

**Supplementary Table 16. Summary statistics of the measures of socio-demographics, lifestyles and developmental adverse events in the baseline.** Med = Median IQR = interquartile range; CV = Coefficient of variation. Under the variable names, there are information about the method to compute these variables and the original variables names in ABCD data dictionary.

| No | Variable | Stats / Values | Graph | Valid |
|---|---|---|---|---|
| 1 | Bilingual Use<br>[numeric]<br>if ACCULT_Q2_Y = 0 then 0<br>otherwise 11-<br>(ACCULT_Q4_Y+ACCULT_Q5_Y) | Mean (sd) : 1 (1.7)<br>min < med < max:<br>0 < 0 < 9<br>IQR (CV) : 1 (1.7) | 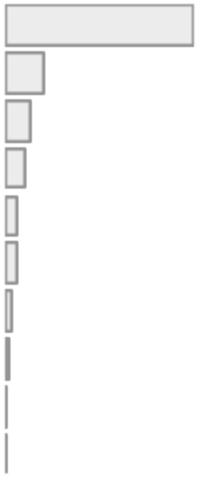 | 11681<br>(99.3%) |
| 2 | Parent Marital Status<br>[factor]<br>DEMO_PRNT_MARITAL_V2 | 1. 1<br>2. 2<br>3. 3<br>4. 4<br>5. 5<br>6. 6 | 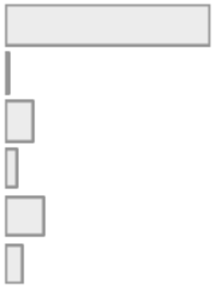 | 11667<br>(99.2%) |
| 3 | Parents' Education<br>[numeric]<br>Mean of<br>(DEMO_PRNT_ED_V2,<br>DEMO_PRTNR_ED_V2) | Mean (sd) : 16.4 (2.7)<br>min < med < max:<br>3 < 17 < 21<br>IQR (CV) : 3.5 (0.2) | 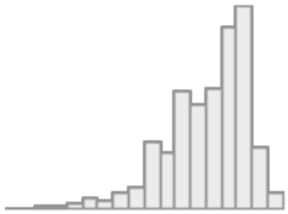 | 11748<br>(99.9%) |
| 4 | Parents' Income<br>[integer]<br>DEMO_COMB_INCOME_V2 | Mean (sd) : 7.2 (2.4)<br>min < med < max:<br>1 < 8 < 10<br>IQR (CV) : 3 (0.3) | 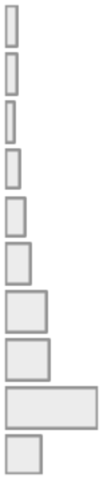 | 10756<br>(91.4%) |
| 5 | Household Size<br>[integer]<br>DEMO_ROSTER_V2 | Mean (sd) : 4.7 (1.6)<br>min < med < max:<br>0 < 4 < 19<br>IQR (CV) : 1 (0.3) | 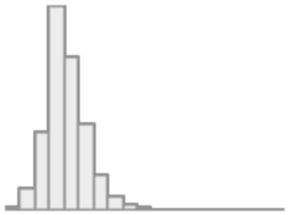 | 11484<br>(97.6%) |

| | | | | |
|---|---|---|---|---|
| 6 | Economics Insecurities<br>[numeric]<br>Sum of<br>(demo_fam_exp1_v2,<br>demo_fam_exp2_v2,<br>demo_fam_exp3_v2,<br>demo_fam_exp4_v2,<br>demo_fam_exp5_v2,<br>demo_fam_exp6_v2,<br>demo_fam_exp7_v2) | Mean (sd) : 0.5 (1.1)<br>min < med < max:<br>0 < 0 < 7<br>IQR (CV) : 0 (2.3) | 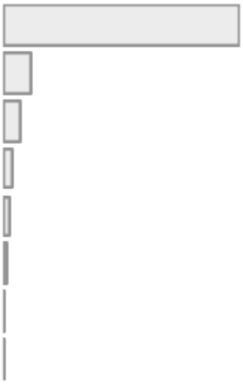 | 11628<br>(98.9%) |
| 7 | Area Deprivation Index<br>[numeric]<br>RESHIST_ADDR1_ADI_WSUM | Mean (sd) : 94.7 (21)<br>min < med < max:<br>1.1 < 98.8 < 125.7<br>IQR (CV) : 20.5 (0.2) | 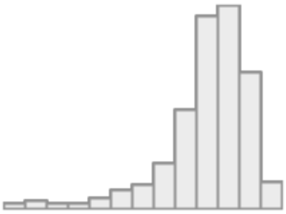 | 10889<br>(92.6%) |
| 8 | Lead Risk<br>[numeric]<br>RESHIST_ADDR1_LEADRISK | Mean (sd) : 5.1 (3.1)<br>min < med < max:<br>1 < 5 < 10<br>IQR (CV) : 6 (0.6) | 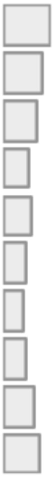 | 11112<br>(94.5%) |
| 9 | Uniform Crime Reports<br>[numeric]<br>(RESHIST_ADDR1_GRNDTOT)/4 | Mean (sd) : 12.1 (5.8)<br>min < med < max:<br>0 < 12.3 < 24.3<br>IQR (CV) : 5.8 (0.5) | 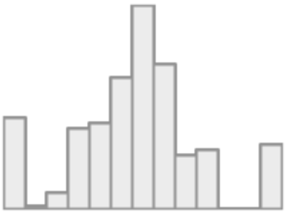 | 11114<br>(94.5%) |
| 10 | Parent reported Neighbourhood Safety<br>[numeric]<br>Sum of<br>(NEIGHBORHOOD1R_P,<br>NEIGHBORHOOD2R_P,<br>NEIGHBORHOOD3R_P) | Mean (sd) : 11.7 (2.9)<br>min < med < max:<br>3 < 12 < 15<br>IQR (CV) : 4 (0.3) | 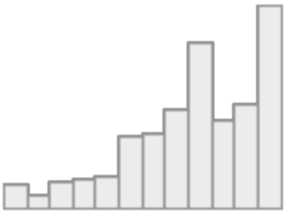 | 11715<br>(99.6%) |
| 11 | Child reported Neighbourhood Safety<br>[numeric]<br>NEIGHBORHOOD_CRIME_Y | Mean (sd) : 4 (1.1)<br>min < med < max:<br>1 < 4 < 5<br>IQR (CV) : 2 (0.3) | 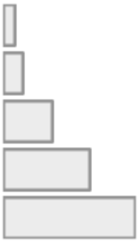 | 11738<br>(99.8%) |

| | | | | |
|---|---|---|---|---|
| 12 | School Environment<br>[numeric]<br>Sum of<br>(SCHOOL_2_Y,<br>SCHOOL_3_Y,<br>SCHOOL_4_Y,<br>SCHOOL_5_Y,<br>SCHOOL_6_Y,<br>SCHOOL_7_Y) | Mean (sd) : 19.9 (2.8)<br>min < med < max:<br>6 < 20 < 24<br>IQR (CV) : 4 (0.1) | 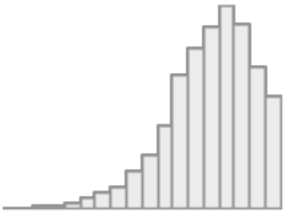 | 11735<br>(99.8%) |
| 13 | School Involvement<br>[numeric]<br>Sum of<br>(SCHOOL_8_Y,<br>SCHOOL_9_Y,<br>SCHOOL_10_Y,<br>SCHOOL_12_Y) | Mean (sd) : 13.1 (2.4)<br>min < med < max:<br>4 < 13 < 16<br>IQR (CV) : 3 (0.2) | 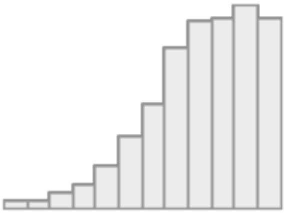 | 11736<br>(99.8%) |
| 14 | School Disengagement<br>[numeric]<br>Sum of<br>(SCHOOL_15_Y,<br>SCHOOL_17_Y) | Mean (sd) : 3.7 (1.5)<br>min < med < max:<br>2 < 4 < 8<br>IQR (CV) : 2 (0.4) | 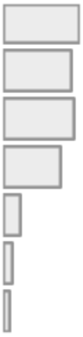 | 11737<br>(99.8%) |
| 15 | Parental Monitoring<br>[numeric]<br>Mean of<br>(PARENT_MONITOR_Q1_Y,<br>PARENT_MONITOR_Q2_Y,<br>PARENT_MONITOR_Q3_Y,<br>PARENT_MONITOR_Q4_Y,<br>PARENT_MONITOR_Q5_Y) | Mean (sd) : 4.4 (0.5)<br>min < med < max:<br>1 < 4.4 < 5<br>IQR (CV) : 0.6 (0.1) | 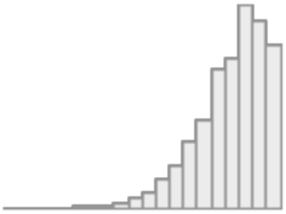 | 11739<br>(99.8%) |
| 16 | Parent reported Family Conflict<br>[numeric]<br>Sum of<br>(FAM_ENVIRO1_P,<br>FAM_ENVIRO2R_P,<br>FAM_ENVIRO3_P,<br>FAM_ENVIRO4R_P,<br>FAM_ENVIRO5_P,<br>FAM_ENVIRO6_P,<br>FAM_ENVIRO7R_P,<br>FAM_ENVIRO8_P,<br>FAM_ENVIRO9R_P) | Mean (sd) : 2.5 (2)<br>min < med < max:<br>0 < 2 < 9<br>IQR (CV) : 3 (0.8) | 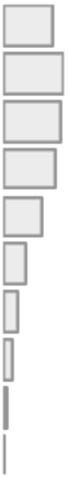 | 11750<br>(99.9%) |

| No | Variable | Stats / Values | Graph | Valid |
|---|---|---|---|---|
| 17 | Child report Family Conflict<br>[numeric]<br>Sum of<br>(FES_YOUTH_Q1,<br>FES_YOUTH_Q2,<br>FES_YOUTH_Q3,<br>FES_YOUTH_Q4,<br>FES_YOUTH_Q5,<br>FES_YOUTH_Q6,<br>FES_YOUTH_Q7,<br>FES_YOUTH_Q8,<br>FES_YOUTH_Q9) | Mean (sd) : 2 (2)<br>min < med < max:<br>0 < 2 < 9<br>IQR (CV) : 3 (1) | 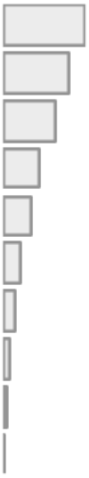 | 11735<br>(99.8%) |
| 18 | Parent reported Prosocial<br>[numeric]<br>Mean of<br>(PROSOCIAL_Q1_P,<br>PROSOCIAL_Q2_P,<br>PROSOCIAL_Q3_P) | Mean (sd) : 1.7 (0.4)<br>min < med < max:<br>0 < 2 < 2<br>IQR (CV) : 0.3 (0.2) | 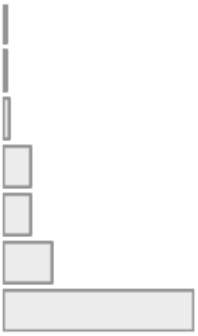 | 11700<br>(99.5%) |
| 19 | Child reported Prosocial<br>[numeric]<br>Mean of<br>(PROSOCIAL_Q1_Y,<br>PROSOCIAL_Q2_Y,<br>PROSOCIAL_Q3_Y) | Mean (sd) : 1.7 (0.4)<br>min < med < max:<br>0 < 1.7 < 2<br>IQR (CV) : 0.7 (0.2) | 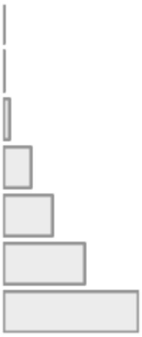 | 11729<br>(99.7%) |
| 20 | Tobbaco Before Pregnant<br>[factor]<br>DEVHX_8_TOBACCO | 1. 0<br>2. 1 | 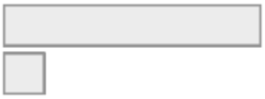 | 11487<br>(97.7%) |
| 21 | Tabbaco After Pregnant<br>[factor]<br>DEVHX_9_TOBACCO | 1. 0<br>2. 1 | 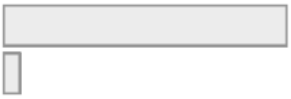 | 11501<br>(97.8%) |
| 22 | Alcohol Before Pregnant<br>[factor]<br>DEVHX_8_ALCOHOL | 1. 0<br>2. 1 | 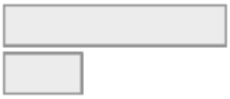 | 11084<br>(94.2%) |
| 23 | Alcohol After Pregnant<br>[factor]<br>DEVHX_9_ALCOHOL | 1. 0<br>2. 1 | 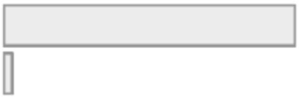 | 11473<br>(97.5%) |
| 24 | Marijuana Before Pregant<br>[factor]<br>DEVHX_8_MARIJUANA | 1. 0<br>2. 1 | 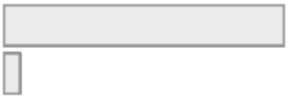 | 11428<br>(97.2%) |
| 25 | Marijuana After Pregant<br>[factor]<br>DEVHX_9_MARIJUANA | 1. 0<br>2. 1 | 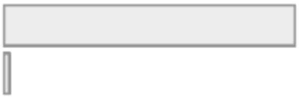 | 11489<br>(97.7%) |

| | | | | |
|---|---|---|---|---|
| 26 | Lack of Sleep<br>[numeric]<br>SLEEPDISTURB1_P | Mean (sd) : 1.7 (0.8)<br>min < med < max:<br>1 < 2 < 5<br>IQR (CV) : 1 (0.5) | 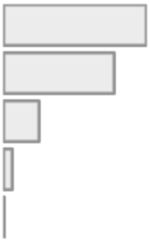 | 11757<br>(100.0%) |
| 27 | Sleep Disturbance<br>[numeric]<br>SLEEPDISTURB2_P | Mean (sd) : 1.9 (1)<br>min < med < max:<br>1 < 2 < 5<br>IQR (CV) : 1 (0.5) | 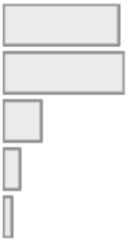 | 11757<br>(100.0%) |
| 28 | Sleep Initiating Maintaining<br>[numeric]<br>SDS_P_SS_DIMS | Mean (sd) : 11.8 (3.7)<br>min < med < max:<br>7 < 11 < 35<br>IQR (CV) : 4 (0.3) | 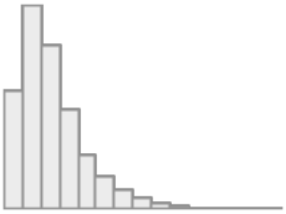 | 11757<br>(100.0%) |
| 29 | Sleep Breathing Disorders<br>[numeric]<br>SDS_P_SS_SBD | Mean (sd) : 3.8 (1.2)<br>min < med < max:<br>3 < 3 < 15<br>IQR (CV) : 1 (0.3) | 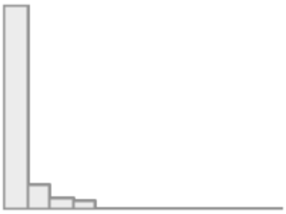 | 11757<br>(100.0%) |
| 30 | Sleep Arousal Disorders<br>[numeric]<br>SDS_P_SS_DA | Mean (sd) : 3.4 (0.9)<br>min < med < max:<br>3 < 3 < 15<br>IQR (CV) : 1 (0.3) | 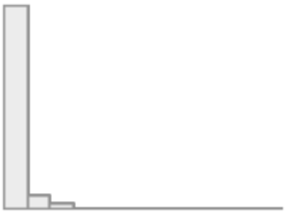 | 11757<br>(100.0%) |
| 31 | Sleep Wake Transition Disorders<br>[numeric]<br>SDS_P_SS_SWTD | Mean (sd) : 8.2 (2.6)<br>min < med < max:<br>6 < 7 < 30<br>IQR (CV) : 3 (0.3) | 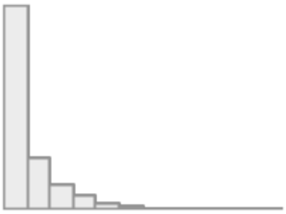 | 11730<br>(99.7%) |
| 32 | Sleep Excessive Somnolence<br>[numeric]<br>SDS_P_SS_DOES | Mean (sd) : 6.9 (2.4)<br>min < med < max:<br>5 < 6 < 25<br>IQR (CV) : 3 (0.4) | 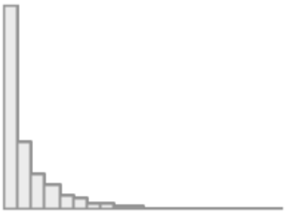 | 11756<br>(99.9%) |

| | | | | |
|---|---|---|---|---|
| 33 | Sleep Hyperhidrosis<br>[numeric]<br>SDS_P_SS_SHY | Mean (sd) : 2.4 (1.2)<br>min < med < max:<br>2 < 2 < 10<br>IQR (CV) : 0 (0.5) | 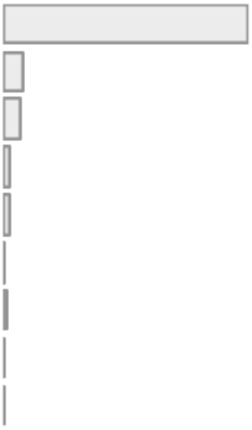 | 11757<br>(100.0%) |
| 34 | Individual Physical Extracurricular Activities<br>[numeric]<br>Baseline sum of<br>(SAI_SS_SBOARD_PERWK_P,<br>SAI_SS_CLIMB_PERWK_P,<br>SAI_SS_GYM_PERWK_P,<br>SAI_SS_ISKATE_PERWK_P,<br>SAI_SS_M_ARTS_PERWK_P,<br>SAI_SS_SKATE_PERWK_P,<br>SAI_SS_DANCE_PERWK_P,<br>SAI_SS_SURF_PERWK_P,<br>SAI_SS_TENNIS_PERWK_P,<br>SAI_SS_RUN_PERWK_P,<br>SAI_SS_MMA_PERWK_P,<br>SAI_SS_YOGA_PERWK_P) | Mean (sd) : 4.9 (5.6)<br>min < med < max:<br>0 < 4 < 70<br>IQR (CV) : 8 (1.2) | 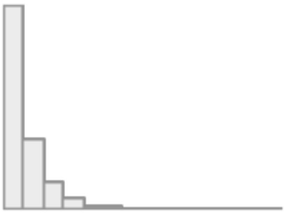 | 11671<br>(99.2%) |
| 35 | Team Physical Extracurricular Activities<br>[numeric]<br>Baseline sum of<br>(SAI_SS_BASE_PERWK_P,<br>SAI_SS_BASKET_PERWK_P,<br>SAI_SS_FHOCK_PERWK_P,<br>SAI_SS_FBALL_PERWK_P,<br>SAI_SS_IHOCK_PERWK_P,<br>SAI_SS_POLO_PERWK_P,<br>SAI_SS_LAX_PERWK_P,<br>SAI_SS_RUGBY_PERWK_P,<br>SAI_SS_SOC_PERWK_P,<br>SAI_SS_WPOLO_PERWK_P,<br>SAI_SS_VBALL_PERWK_P) | Mean (sd) : 8.1 (7.6)<br>min < med < max:<br>0 < 6 < 54<br>IQR (CV) : 13 (0.9) | 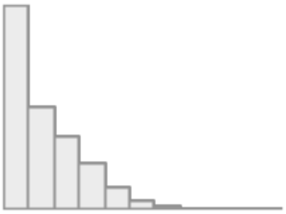 | 11653<br>(99.1%) |
| 36 | Non Physical Extracurricular Activities<br>[numeric]<br>Baseline sum of<br>(SAI_SS_COLLECT_PERWK_P,<br>SAI_SS_MUSIC_PERWK_P,<br>SAI_SS_ART_PERWK_P,<br>SAI_SS_DRAMA_PERWK_P,<br>SAI_SS_CRAFTS_PERWK_P,<br>SAI_SS_CHESS_PERWK_P) | Mean (sd) : 4.9 (6.2)<br>min < med < max:<br>0 < 4 < 54<br>IQR (CV) : 8 (1.3) | 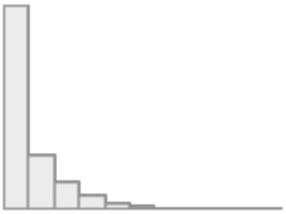 | 11646<br>(99.0%) |

| | | | | |
|---|---|---|---|---|
| 37 | Physically Active<br>[numeric]<br>PHYSICAL_ACTIVITY1_Y | Mean (sd) : 3.5 (2.3)<br>min < med < max:<br>0 < 3 < 7<br>IQR (CV) : 3 (0.7) | 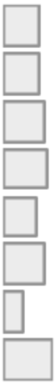 | 11734<br>(99.8%) |
| 38 | Mature Video Games Play<br>[numeric]<br>SCREEN13_Y | Mean (sd) : 0.6 (0.9)<br>min < med < max:<br>0 < 0 < 3<br>IQR (CV) : 1 (1.5) | 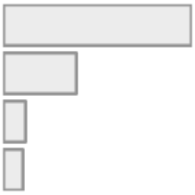 | 11742<br>(99.8%) |
| 39 | Mature Movies Watch<br>[numeric]<br>SCREEN14_Y | Mean (sd) : 0.4 (0.6)<br>min < med < max:<br>0 < 0 < 3<br>IQR (CV) : 1 (1.7) | 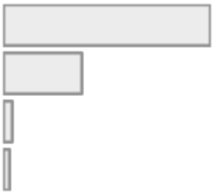 | 11741<br>(99.8%) |
| 40 | Weekday Screen Use<br>[numeric]<br>Sum of<br>(SCREEN1_WKDY_Y,<br>SCREEN2_WKDY_Y,<br>SCREEN3_WKDY_Y,<br>SCREEN4_WKDY_Y,<br>SCREEN5_WKDY_Y,<br>SCREEN_WKDY_Y) | Mean (sd) : 3.5 (3.1)<br>min < med < max:<br>0 < 2.5 < 24<br>IQR (CV) : 3.5 (0.9) | 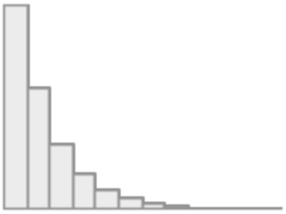 | 11725<br>(99.7%) |
| 41 | Weekend Screen Use<br>[numeric]<br>Sum of<br>(SCREEN7_WKND_Y,<br>SCREEN8_WKND_Y,<br>SCREEN9_WKND_Y,<br>SCREEN10_WKND_Y,<br>SCREEN11_WKND_Y,<br>SCREEN12_WKND_Y) | Mean (sd) : 4.6 (3.6)<br>min < med < max:<br>0 < 3.5 < 24<br>IQR (CV) : 4.2 (0.8) | 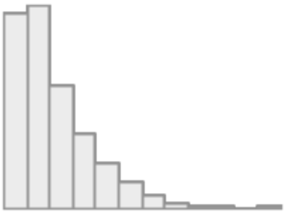 | 11720<br>(99.6%) |
| 42 | Developmental Prematurity<br>[factor]<br>DEVHX_12A_P | 1. 0<br>2. 1 | 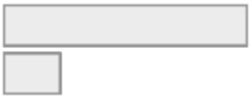 | 11618<br>(98.8%) |

| | | | | |
|---|---|---|---|---|
| 43 | Birth Complications<br>[numeric]<br>Sum of<br>(DEVHX_14A3_P,<br>DEVHX_14B3_P,<br>DEVHX_14C3_P,<br>DEVHX_14D3_P,<br>DEVHX_14E3_P,<br>DEVHX_14F3_P,<br>DEVHX_14G3_P,<br>DEVHX_14H3_P) | Mean (sd) : 0.4 (0.7)<br>min < med < max:<br>0 < 0 < 8<br>IQR (CV) : 1 (2) | 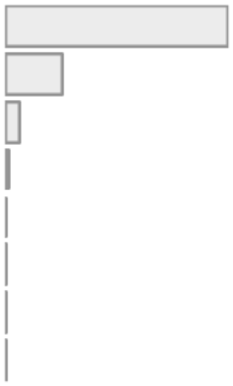 | 11007<br>(93.6%) |
| 44 | Pregnancy Complications<br>[numeric]<br>Sum of<br>(DEVHX_10A3_P,<br>DEVHX_10B3_P,<br>DEVHX_10C3_P,<br>DEVHX_10D3_P,<br>DEVHX_10E3_P,<br>DEVHX_10F3_P,<br>DEVHX_10G3_P,<br>DEVHX_10H3_P,<br>DEVHX_10I3_P,<br>DEVHX_10J3_P,<br>DEVHX_10K3_P,<br>DEVHX_10L3_P) | Mean (sd) : 0.6 (1)<br>min < med < max:<br>0 < 0 < 12<br>IQR (CV) : 1 (1.7) | 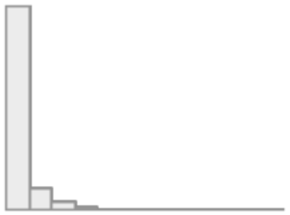 | 11027<br>(93.8%) |

**Supplementary Table 17. Summary statistics of the measures of socio-demographics, lifestyles and developmental adverse events in the follow up.** We only provided variables that were repeatedly correction in the follow up here. Med = Median IQR = interquartile range; CV = Coefficient of variation. Under the variable names, there are information about the method to compute these variables and the original variables names in ABCD data dictionary.

| No | Variable | Stats / Values | Graph | Valid |
|---|---|---|---|---|
| 1 | Bilingual Use<br>[numeric]<br>if ACCULT_Q2_Y = 0 then 0<br>otherwise 11-<br>(ACCULT_Q4_Y+ACCULT_Q5_Y) | Mean (sd) : 1 (1.6)<br>min < med < max:<br>0 < 0 < 9<br>IQR (CV) : 1 (1.6) | 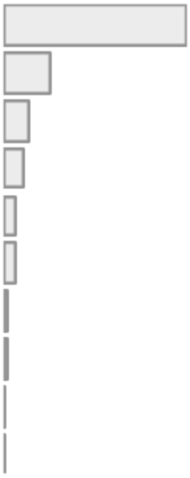 | 10768<br>(99.3%) |
| 2 | Parent Marital Status<br>[factor]<br>DEMO_PRNT_MARITAL_V2_L | 1. 1<br>2. 2<br>3. 3<br>4. 4<br>5. 5<br>6. 6 | 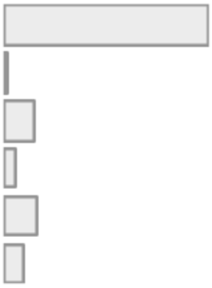 | 10766<br>(99.3%) |
| 3 | Parents' Education<br>[numeric]<br>Mean of<br>(DEMO_PRNT_ED_V2_2YR_L,<br>DEMO_PRTNR_ED_V2_2YR_L) | Mean (sd) : 17.7 (3.1)<br>min < med < max:<br>1 < 18 < 23<br>IQR (CV) : 3 (0.2) | 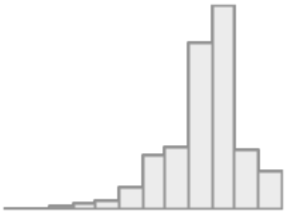 | 10823<br>(99.8%) |
| 4 | Parents' Income<br>[integer]<br>DEMO_COMB_INCOME_V2_L | Mean (sd) : 7.5 (2.3)<br>min < med < max:<br>1 < 8 < 10<br>IQR (CV) : 2 (0.3) | 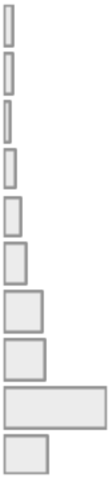 | 9953<br>(91.8%) |

| No | Variable | Stats / Values | Graph | Valid |
|---|---|---|---|---|
| 5 | Economics Insecurities<br>[numeric]<br>sum of<br>(demo_fam_exp1_v2_l,<br>demo_fam_exp2_v2_l,<br>demo_fam_exp3_v2_l,<br>demo_fam_exp4_v2_l,<br>demo_fam_exp5_v2_l,<br>demo_fam_exp6_v2_l,<br>demo_fam_exp7_v2_l) | Mean (sd) : 0.4 (1)<br>min < med < max:<br>0 < 0 < 7<br>IQR (CV) : 0 (2.5) | 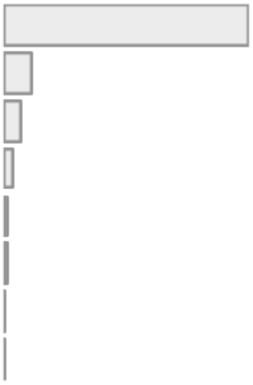 | 10734<br>(99.0%) |
| 6 | Parent reported Neighbourhood Safety<br>[numeric]<br>Sum of<br>(NEIGHBORHOOD1R_P,<br>NEIGHBORHOOD2R_P,<br>NEIGHBORHOOD3R_P) | Mean (sd) : 11.6 (2.8)<br>min < med < max:<br>3 < 12 < 15<br>IQR (CV) : 4 (0.2) | 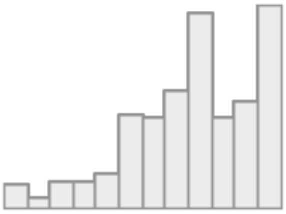 | 10752<br>(99.2%) |
| 7 | Child reported Neighbourhood Safety<br>[numeric]<br>NEIGHBORHOOD_CRIME_Y | Mean (sd) : 4.1 (1)<br>min < med < max:<br>1 < 4 < 5<br>IQR (CV) : 1 (0.2) | 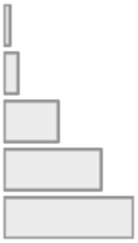 | 10811<br>(99.7%) |
| 8 | School Environment<br>[numeric]<br>Sum of<br>(SCHOOL_2_Y,<br>SCHOOL_3_Y,<br>SCHOOL_4_Y,<br>SCHOOL_5_Y,<br>SCHOOL_6_Y,<br>SCHOOL_7_Y) | Mean (sd) : 19.6 (2.8)<br>min < med < max:<br>6 < 20 < 24<br>IQR (CV) : 4 (0.1) | 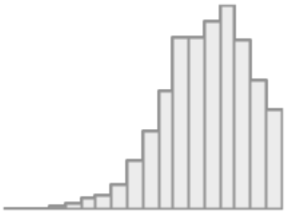 | 10808<br>(99.7%) |
| 9 | School Involvement<br>[numeric]<br>Sum of<br>(SCHOOL_8_Y,<br>SCHOOL_9_Y,<br>SCHOOL_10_Y,<br>SCHOOL_12_Y) | Mean (sd) : 12.6 (2.3)<br>min < med < max:<br>4 < 13 < 16<br>IQR (CV) : 3 (0.2) | 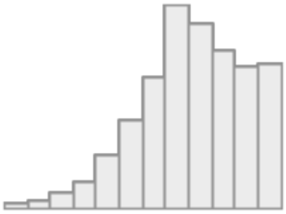 | 10808<br>(99.7%) |
| 10 | School Disengagement<br>[numeric]<br>Sum of<br>(SCHOOL_15_Y,<br>SCHOOL_17_Y) | Mean (sd) : 4 (1.3)<br>min < med < max:<br>2 < 4 < 8<br>IQR (CV) : 2 (0.3) | 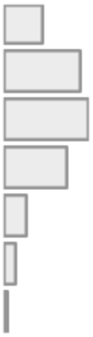 | 10808<br>(99.7%) |

| No | Variable | Stats / Values | Graph | Valid |
|---|---|---|---|---|
| 11 | Parental Monitoring<br>[numeric]<br>Mean of<br>(PARENT_MONITOR_Q1_Y,<br>PARENT_MONITOR_Q2_Y,<br>PARENT_MONITOR_Q3_Y,<br>PARENT_MONITOR_Q4_Y,<br>PARENT_MONITOR_Q5_Y) | Mean (sd) : 4.5 (0.5)<br>min < med < max:<br>1 < 4.6 < 5<br>IQR (CV) : 0.6 (0.1) | 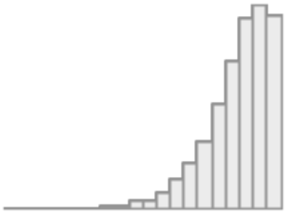 | 10811<br>(99.7%) |
| 12 | Parent reported Family Conflict<br>[numeric]<br>Sum of<br>(FAM_ENVIRO1_P,<br>FAM_ENVIRO2R_P,<br>FAM_ENVIRO3_P,<br>FAM_ENVIRO4R_P,<br>FAM_ENVIRO5_P,<br>FAM_ENVIRO6_P,<br>FAM_ENVIRO7R_P,<br>FAM_ENVIRO8_P,<br>FAM_ENVIRO9R_P) | Mean (sd) : 2.4 (2)<br>min < med < max:<br>0 < 2 < 9<br>IQR (CV) : 3 (0.8) | 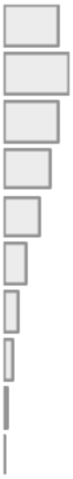 | 10813<br>(99.8%) |
| 13 | Child report Family Conflict<br>[numeric]<br>Sum of<br>(FES_YOUTH_Q1,<br>FES_YOUTH_Q2,<br>FES_YOUTH_Q3,<br>FES_YOUTH_Q4,<br>FES_YOUTH_Q5,<br>FES_YOUTH_Q6,<br>FES_YOUTH_Q7,<br>FES_YOUTH_Q8,<br>FES_YOUTH_Q9) | Mean (sd) : 1.9 (1.8)<br>min < med < max:<br>0 < 1 < 9<br>IQR (CV) : 3 (1) | 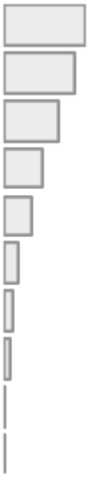 | 10268<br>(94.7%) |
| 14 | Parent reported Prosocial<br>[numeric]<br>Mean of<br>(PROSOCIAL_Q1_P,<br>PROSOCIAL_Q2_P,<br>PROSOCIAL_Q3_P) | Mean (sd) : 1.7 (0.4)<br>min < med < max:<br>0 < 2 < 2<br>IQR (CV) : 0.3 (0.2) | 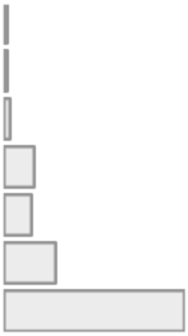 | 10788<br>(99.5%) |

| No | Variable | Stats / Values | Graph | Valid |
|---|---|---|---|---|
| 15 | Child reported Prosocial<br>[numeric]<br>Mean of<br>(PROSOCIAL_Q1_Y,<br>PROSOCIAL_Q2_Y,<br>PROSOCIAL_Q3_Y) | Mean (sd) : 1.7 (0.4)<br>min < med < max:<br>0 < 2 < 2<br>IQR (CV) : 0.7 (0.2) | 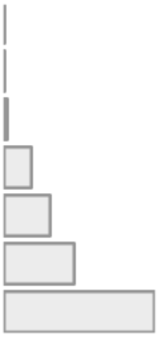 | 10811<br>(99.7%) |
| 16 | Lack of Sleep<br>[numeric]<br>SLEEPDISTURB1_P | Mean (sd) : 2 (0.9)<br>min < med < max:<br>1 < 2 < 5<br>IQR (CV) : 1 (0.4) | 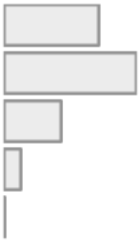 | 10803<br>(99.7%) |
| 17 | Sleep Disturbance<br>[numeric]<br>SLEEPDISTURB2_P | Mean (sd) : 2.1 (1.1)<br>min < med < max:<br>1 < 2 < 5<br>IQR (CV) : 2 (0.5) | 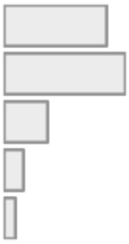 | 10803<br>(99.7%) |
| 18 | Sleep Initiating Maintaining<br>[numeric]<br>SDS_P_SS_DIMS | Mean (sd) : 12 (3.8)<br>min < med < max:<br>7 < 11 < 34<br>IQR (CV) : 5 (0.3) | 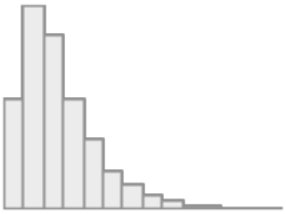 | 10803<br>(99.7%) |
| 19 | Sleep Breathing Disorders<br>[numeric]<br>SDS_P_SS_SBD | Mean (sd) : 3.7 (1.2)<br>min < med < max:<br>3 < 3 < 15<br>IQR (CV) : 1 (0.3) | 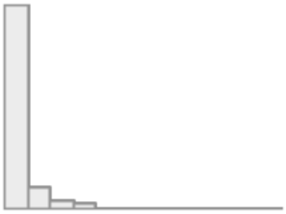 | 10803<br>(99.7%) |
| 20 | Sleep Arousal Disorders<br>[numeric]<br>SDS_P_SS_DA | Mean (sd) : 3.3 (0.7)<br>min < med < max:<br>3 < 3 < 11<br>IQR (CV) : 0 (0.2) | 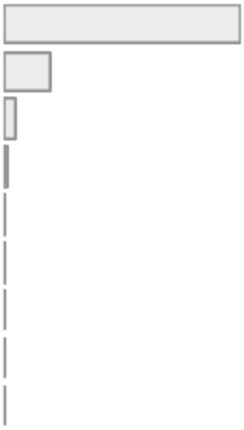 | 10803<br>(99.7%) |

| No | Variable | Stats / Values | Graph | Valid |
|---|---|---|---|---|
| 21 | Sleep Wake Transition Disorders<br>[numeric]<br>SDS_P_SS_SWTD | Mean (sd) : 7.8 (2.4)<br>min < med < max:<br>6 < 7 < 28<br>IQR (CV) : 3 (0.3) | 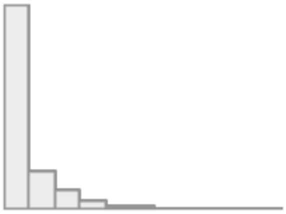 | 10802<br>(99.6%) |
| 22 | Sleep Excessive Somnolence<br>[numeric]<br>SDS_P_SS_DOES | Mean (sd) : 7.1 (2.6)<br>min < med < max:<br>5 < 6 < 25<br>IQR (CV) : 3 (0.4) | 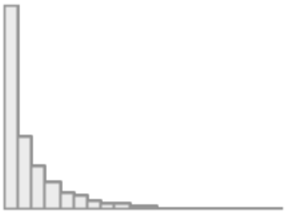 | 10803<br>(99.7%) |
| 23 | Sleep Hyperhidrosis<br>[numeric]<br>SDS_P_SS_SHY | Mean (sd) : 2.3 (1)<br>min < med < max:<br>2 < 2 < 10<br>IQR (CV) : 0 (0.4) | 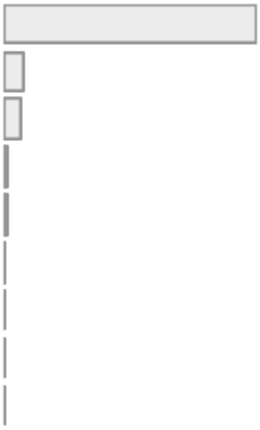 | 10803<br>(99.7%) |
| 24 | Individual Physical Extracurricular Activities<br>[numeric]<br>Summation of<br>(SAI_SS_SBOARD_PERWK_P,<br>SAI_SS_CLIMB_PERWK_P,<br>SAI_SS_GYM_PERWK_P,<br>SAI_SS_ISKATE_PERWK_P,<br>SAI_SS_M_ARTS_PERWK_P,<br>SAI_SS_SKATE_PERWK_P,<br>SAI_SS_DANCE_PERWK_P,<br>SAI_SS_SURF_PERWK_P,<br>SAI_SS_TENNIS_PERWK_P,<br>SAI_SS_RUN_PERWK_P,<br>SAI_SS_MMA_PERWK_P,<br>SAI_SS_YOGA_PERWK_P) in the baseline, and<br>(SAI_SS_SBOARD_PERWK_P_L,<br>SAI_SS_CLIMB_PERWK_P_L,<br>SAI_SS_GYM_PERWK_P_L,<br>SAI_SS_ISKATE_PERWK_P_L,<br>SAI_SS_M_ARTS_PERWK_P_L,<br>SAI_SS_SKATE_PERWK_P_L,<br>SAI_SS_DANCE_PERWK_P_L,<br>SAI_SS_SURF_PERWK_P_L,<br>SAI_SS_TENNIS_PERWK_P_L,<br>SAI_SS_RUN_PERWK_P_L,<br>SAI_SS_MMA_PERWK_P_L, | Mean (sd) : 9.8 (10.5)<br>min < med < max:<br>0 < 7 < 107<br>IQR (CV) : 16 (1.1) | 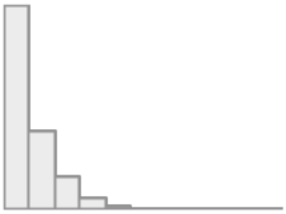 | 10416<br>(96.1%) |

| No | Variable | Stats / Values | Graph | Valid |
|---|---|---|---|---|
| | SAI_SS_YOGA_PERWK_P_L) in the one-year follow-up, and<br>(SAI_SS_SBOARD_PERWK_P_L,<br>SAI_SS_CLIMB_PERWK_P_L,<br>SAI_SS_GYM_PERWK_P_L,<br>SAI_SS_ISKATE_PERWK_P_L,<br>SAI_SS_M_ARTS_PERWK_P_L,<br>SAI_SS_SKATE_PERWK_P_L ,<br>SAI_SS_DANCE_PERWK_P_L,<br>SAI_SS_SURF_PERWK_P_L ,<br>SAI_SS_TENNIS_PERWK_P_L,<br>SAI_SS_RUN_PERWK_P_L,<br>SAI_SS_MMA_PERWK_P_L,<br>SAI_SS_YOGA_PERWK_P_L) in the two-year follow-up. | | | |
| 25 | Team Physical Extracurricular Activities<br>[numeric]<br>summation of<br>(SAI_SS_BASE_PERWK_P,<br>SAI_SS_BASKET_PERWK_P,<br>SAI_SS_FHOCK_PERWK_P,<br>SAI_SS_FBALL_PERWK_P,<br>SAI_SS_IHOCK_PERWK_P,<br>SAI_SS_POLO_PERWK_P,<br>SAI_SS_LAX_PERWK_P,<br>SAI_SS_RUGBY_PERWK_P,<br>SAI_SS_SOC_PERWK_P,<br>SAI_SS_WPOLO_PERWK_P,<br>SAI_SS_VBALL_PERWK_P) in the baseline, and<br>(SAI_SS_BASE_PERWK_P_L,<br>SAI_SS_BASKET_PERWK_P_L,<br>SAI_SS_FHOCK_PERWK_P_L,<br>SAI_SS_FBALL_PERWK_P_L,<br>SAI_SS_IHOCK_PERWK_P_L,<br>SAI_SS_POLO_PERWK_P_L,<br>SAI_SS_LAX_PERWK_P_L,<br>SAI_SS_RUGBY_PERWK_P_L,<br>SAI_SS_SOC_PERWK_P_L,<br>SAI_SS_WPOLO_PERWK_P_L,<br>SAI_SS_VBALL_PERWK_P_L<br>) in the one-year follow-up, and<br>( SAI_SS_BASE_PERWK_P_L,<br>SAI_SS_BASKET_PERWK_P_L,<br>SAI_SS_FHOCK_PERWK_P_L,<br>SAI_SS_FBALL_PERWK_P_L,<br>SAI_SS_IHOCK_PERWK_P_L,<br>SAI_SS_POLO_PERWK_P_L,<br>SAI_SS_LAX_PERWK_P_L,<br>SAI_SS_RUGBY_PERWK_P_L,<br>SAI_SS_SOC_PERWK_P_L,<br>SAI_SS_WPOLO_PERWK_P_L,<br>SAI_SS_VBALL_PERWK_P_L)<br>) in the two-year follow-up | Mean (sd) : 18.2 (16.5)<br>min < med < max:<br>0 < 15 < 90<br>IQR (CV) : 23 (0.9) | 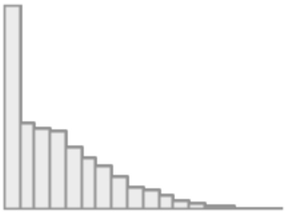 | 10405<br>(96.0%) |

| No | Variable | Stats / Values | Graph | Valid |
|---|---|---|---|---|
| 26 | Non Physical Extracurricular Activities<br>[numeric]<br>Summation of (SAI_SS_COLLECT_PERWK_P, SAI_SS_MUSIC_PERWK_P, SAI_SS_ART_PERWK_P, SAI_SS_DRAMA_PERWK_P, SAI_SS_CRAFTS_PERWK_P, SAI_SS_CHESS_PERWK_P) in the baseline and (SAI_SS_COLLECT_PERWK_P_L, SAI_SS_MUSIC_PERWK_P_L, SAI_SS_ART_PERWK_P_L, SAI_SS_DRAMA_PERWK_P_L, SAI_SS_CRAFTS_PERWK_P_L, SAI_SS_CHESS_PERWK_P_L) in the One-year follow-up and (SAI_SS_COLLECT_PERWK_P_L, SAI_SS_MUSIC_PERWK_P_L, SAI_SS_ART_PERWK_P_L, SAI_SS_DRAMA_PERWK_P_L, SAI_SS_CRAFTS_PERWK_P_L, SAI_SS_CHESS_PERWK_P_L) in the two-year follow-up) | Mean (sd) : 14 (14)<br>min < med < max:<br>0 < 11 < 90<br>IQR (CV) : 21 (1) | 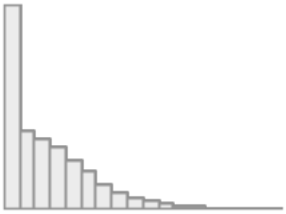 | 10390<br>(95.8%) |
| 27 | Physically Active<br>[numeric]<br>PHYSICAL_ACTIVITY1_Y | Mean (sd) : 3.8 (2.2)<br>min < med < max:<br>0 < 4 < 7<br>IQR (CV) : 3 (0.6) | 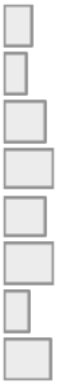 | 10813<br>(99.8%) |
| 28 | Mature Video Games Play<br>[numeric]<br>SCREEN13_Y | Mean (sd) : 0.6 (0.9)<br>min < med < max:<br>0 < 0 < 3<br>IQR (CV) : 1 (1.4) | 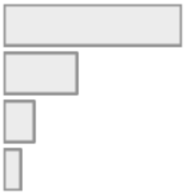 | 10812<br>(99.7%) |
| 29 | Mature Movies Watch<br>[numeric]<br>SCREEN14_Y | Mean (sd) : 0.5 (0.7)<br>min < med < max:<br>0 < 0 < 3<br>IQR (CV) : 1 (1.3) | 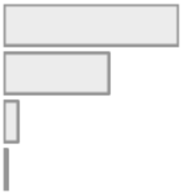 | 10804<br>(99.7%) |

**Supplementary Table 18. The differences in social demographics, lifestyles and developmental adverse events between participants who provided cognitive scores in the follow up.** We used social demographics, lifestyles and developmental adverse events collected at baseline.

| Variable Names | Having cognitive scores in the follow up. | Not having cognitive scores in the follow up. | Test statistics |
|---|---|---|---|
| Age in months | Mean (sd) : 119.3 (7.5) | Mean (sd) : 118.3 (7.6) | Yuen's $t(3783) = 6.05$, $p < 0.001$, Cohen's $d = 0.092$ |
| Sex | Male = 3918 (52.4%)<br>Female = 3564 (47.6%)<br>Intersex-Male = 1 (0.0%)<br>Intersex-female = 0 (0.0%)<br>Do not know = 0 (0.0%) | Male = 1776(53.2%)<br>Female = 1563(46.8%)<br>Intersex-Male = 2(0.1%)<br>Intersex-female = 0(0.0%)<br>Do not know = 0 (0.0%) | $\chi^2$ (4, $N$ = 10824) = 6, $p = 0.199$ |
| Body Mass Index | Mean (sd) : 18.7 (4.1) | Mean (sd) : 18.9 (4.4) | Yuen's $t$ (3658) = 1.605, $p = 0.109$, Cohen's $d = 0.023$ |
| Race | White = 4190(56.0%)<br>Black = 918(12.3%)<br>Hispanic = 1441(19.3%)<br>Asian = 157(2.1%)<br>Other = 777(10.4%) | White = 1611(48.2%)<br>Black = 612(18.3%)<br>Hispanic = 689(20.6%)<br>Asian = 68(2.0%)<br>Other = 360(10.8%) | $\chi^2$ (16, $N$ = 10823) = 20, $p = 0.22$ |
| Bilingual Use | Mean (sd) : 1 (1.7) | Mean (sd) : 1 (1.7) | Yuen's $t$ (3776) = 0.696, $p = 0.486$, Cohen's $d = 0.011$ |
| Parent Marital Status | Married = 5239(70.5%)<br>Widowed = 59(0.8%)<br>Divorced = 684(9.2%)<br>Separated = 264(3.6%)<br>NeverMarried = 806(10.8%)<br>LivingWithPartner =381(5.1%) | Married = 2194(66.0%)<br>Widowed = 29(0.9%)<br>Divorced =290(8.7%)<br>Separated = 135(4.1%)<br>NeverMarried = 460(13.8%)<br>LivingWithPartner =214(6.4%) | $\chi^2$ (25, $N$ = 10755) = 30, $p = 0.224$ |
| Parents' Education | Mean (sd) : 16.6 (2.6) | Mean (sd) : 16.3 (2.8) | Yuen's $t$ (3262) = 4.175, $p$ <0.001, Cohen's $d = 0.068$ |
| Parents' Income | Mean (sd) : 7.4 (2.3) | Mean (sd) : 7.2 (2.5) | Yuen's $t$ (2854) = 2.243, $p = 0.025$, Cohen's $d = 0.034$ |
| Household Size | Mean (sd) : 4.7 (1.5) | Mean (sd) : 4.7 (1.6) | Yuen's $t$ (3718) = 0.39, $p = 0.697$, Cohen's $d = 0.007$ |
| Economics Insecurities | Mean (sd) : 0.4 (1.1) | Mean (sd) : 0.5 (1.1) | Yuen's $t$ (1982) = 2.65, $p = 0.008$, Cohen's $d = 0.033$ |
| Area Deprivation Index | Mean (sd) : 94.6 (20.7) | Mean (sd) : 94.9 (21.2) | Yuen's $t$ (3297) = 1.686, $p = 0.092$, Cohen's $d = 0.029$ |
| Lead Risk | Mean (sd) : 5 (3.1) | Mean (sd) : 5.1 (3.1) | Yuen's $t$ (3374) = 1.797, $p = 0.072$, Cohen's $d = 0.027$ |
| Uniform Crime Reports | Mean (sd) : 12.1 (5.5) | Mean (sd) : 12 (6.1) | Yuen's $t$ (3370) = 0.873, $p = 0.383$, Cohen's $d = 0.014$ |
| Parent reported Neighbourhood Safety | Mean (sd) : 11.8 (2.9) | Mean (sd) : 11.6 (3) | Yuen's $t$ (3382) = 1.799, $p = 0.072$, Cohen's $d = 0.025$ |
| Child reported Neighbourhood Safety | Mean (sd) : 4.1 (1.1) | Mean (sd) : 4 (1.1) | Yuen's $t$ (3786) = 2.258, $p = 0.024$, Cohen's $d = 0.036$ |
| School Environment | Mean (sd) : 20 (2.8) | Mean (sd) : 19.8 (2.9) | Yuen's $t$ (3787) = 1.763, $p = 0.078$, Cohen's $d = 0.029$ |
| School Involvement | Mean (sd) : 13.1 (2.3) | Mean (sd) : 12.9 (2.4) | Yuen's $t$ (3790) = 3.203, $p = 0.001$, Cohen's $d = 0.05$ |
| School Disengagement | Mean (sd) : 3.7 (1.4) | Mean (sd) : 3.8 (1.5) | Yuen's $t$ (3800) = 2.171, $p = 0.03$, Cohen's $d = 0.035$ |
| Lack of Sleep | Mean (sd) : 1.7 (0.8) | Mean (sd) : 1.7 (0.8) | Yuen's $t$ (3860) = 3.084, $p = 0.002$, Cohen's $d = 0.05$ |
| Sleep Disturbance | Mean (sd) : 1.9 (1) | Mean (sd) : 1.9 (1) | Yuen's $t$ (3877) = 1.567, $p = 0.117$, Cohen's $d = 0.025$ |
| Sleep Initiating Maintaining | Mean (sd) : 11.7 (3.7) | Mean (sd) : 11.9 (3.8) | Yuen's $t$ (3862) = 2.481, $p = 0.013$, Cohen's $d = 0.038$ |
| Sleep Breathing Disorders | Mean (sd) : 3.7 (1.2) | Mean (sd) : 3.8 (1.3) | Yuen's $t$ (3834) = 1.43, $p = 0.153$, Cohen's $d = 0.022$ |
| Sleep Arousal Disorders | Mean (sd) : 3.4 (0.9) | Mean (sd) : 3.4 (1) | Yuen's $t$ (3885) = 0.966, $p = 0.334$, Cohen's $d = 0.013$ |
| Sleep Wake Transition Disorders | Mean (sd) : 8.2 (2.6) | Mean (sd) : 8.1 (2.6) | Yuen's $t$ (3828) = 1.198, $p = 0.231$, Cohen's $d = 0.022$ |
| Sleep Excessive Somnolence | Mean (sd) : 6.9 (2.4) | Mean (sd) : 7 (2.5) | Yuen's $t$ (3836) = 0.131, $p = 0.896$, Cohen's $d = 0.007$ |
| Sleep Hyperhidrosis | Mean (sd) : 2.4 (1.2) | Mean (sd) : 2.5 (1.2) | Yuen's $t$ (4375) = 1.755, $p = 0.079$, Cohen's $d = 0.029$ |
| Individual Physical Extracurricular Activities | Mean (sd) : 5 (5.7) | Mean (sd) : 4.7 (5.4) | Yuen's $t$ (4173) = 2.933, $p = 0.003$, Cohen's $d = 0.044$ |
| Team Physical Extracurricular Activities | Mean (sd) : 8.4 (7.7) | Mean (sd) : 7.8 (7.4) | Yuen's $t$ (4007) = 3.604, $p$ <0.001, Cohen's $d = 0.055$ |
| Non Physical Extracurricular Activities | Mean (sd) : 5.1 (6.3) | Mean (sd) : 4.8 (6.1) | Yuen's $t$ (4075) = 2.961, $p = 0.003$, Cohen's $d = 0.047$ |
| Physically Active | Mean (sd) : 3.5 (2.3) | Mean (sd) : 3.4 (2.3) | Yuen's $t$ (3838) = 2.094, $p = 0.036$, Cohen's $d = 0.033$ |
| Mature Video Games Play | Mean (sd) : 0.5 (0.8) | Mean (sd) : 0.6 (0.9) | Yuen's $t$ (3816) = 1.396, $p = 0.163$, Cohen's $d = 0.022$ |
| Mature Movies Watch | Mean (sd) : 0.4 (0.6) | Mean (sd) : 0.4 (0.7) | Yuen's $t$ (3728) = 4.038, |

| | | | |
|---|---|---|---|
| | | | $p$ <0.001, Cohen's $d$ = 0.065 |
| Weekday Screen Use | Mean (sd) : 3.3 (3) | Mean (sd) : 3.6 (3.3) | Yuen's $t$ (3220) = 4.161, $p$ <0.001, Cohen's $d$ = 0.069 |
| Weekend Screen Use | Mean (sd) : 4.5 (3.5) | Mean (sd) : 4.8 (3.7) | Yuen's $t$ (3521) = 3.218, $p$ = 0.001, Cohen's $d$ = 0.053 |
| Tobacco Before Pregnant | No = 6328(86.7%)<br>Yes = 974(13.3%) | No = 2838(86.7%)<br>Yes = 436(13.3%) | $\chi^2$ (1, $N$ = 10576) = 0, p = 1 |
| Tobacco After Pregnant | No = 6968(95.2%)<br>Yes = 351(4.8%) | No = 3081(94.2%)<br>Yes = 190(5.8%) | $\chi^2$ (1, $N$ = 10590) = 0, p = 1 |
| Alcohol Before Pregnant | No = 5174(73.4%)<br>Yes = 1871(26.6%) | No = 2380(75.4%)<br>Yes = 775(24.6%) | $\chi^2$ (1, $N$ = 10200) = 0, p = 1 |
| Alcohol After Pregnant | No = 7096(97.1%)<br>Yes = 210(2.9%) | No = 3175(97.4%)<br>Yes = 85(2.6%) | $\chi^2$ (1, $N$ = 10566) = 0, p = 1 |
| Marijuana Before Pregnant | No = 6874(94.5%)<br>Yes = 399(5.5%) | No = 3044(93.9%)<br>Yes = 199(6.1%) | $\chi^2$ (1, $N$ = 10516) = 0, p = 1 |
| Marijuana After Pregnant | No = 7182(98.2%)<br>Yes = 130(1.8%) | No = 3191(97.7%)<br>Yes = 74(2.3%) | $\chi^2$ (1, $N$ = 10577) = 0, p = 1 |
| Developmental Prematurity | No = 5945(80.3%)<br>Yes = 1458(19.7%) | No = 2735(83.0%)<br>Yes = 561(17.0%) | $\chi^2$ (1, $N$ = 10699) = 0, p = 1 |
| Birth Complications | Mean (sd) : 0.4 (0.8) | Mean (sd) : 0.4 (0.7) | Yuen's $t$ (3591) = 0.121, $p$ = 0.904, Cohen's $d$ = 0.007 |
| Pregnancy Complications | Mean (sd) : 0.6 (1) | Mean (sd) : 0.6 (1) | Yuen's $t$ (3543) = 1.19, $p$ = 0.234, Cohen's $d$ = 0.018 |
| Parental Monitoring | Mean (sd) : 4.4 (0.5) | Mean (sd) : 4.4 (0.5) | Yuen's $t$ (3810) = 0.451, $p$ = 0.652, Cohen's $d$ = 0.009 |
| Parent reported Family Conflict | Mean (sd) : 2.5 (1.9) | Mean (sd) : 2.6 (2) | Yuen's $t$ (3805) = 1.404, $p$ = 0.16, Cohen's $d$ = 0.023 |
| Child report Family Conflict | Mean (sd) : 2 (1.9) | Mean (sd) : 2.1 (2) | Yuen's $t$ (3809) = 1.751, $p$ = 0.08, Cohen's $d$ = 0.026 |
| Parent reported Prosocial | Mean (sd) : 1.8 (0.4) | Mean (sd) : 1.8 (0.4) | Yuen's $t$ (3817) = 0.288, $p$ = 0.774, Cohen's $d$ = 0.007 |
| Child reported Prosocial | Mean (sd) : 1.7 (0.4) | Mean (sd) : 1.7 (0.4) | Yuen's $t$ (3849) = 2.529, $p$ = 0.011, Cohen's $d$ = 0.041 |

**Supplementary Table 19. Medication reports in the baseline and follow up.** This report is derived from the su_y_plus table and utilises the Anatomical Therapeutic Chemical (ATC) Classification System for to group medications according to their functionality.

| Functionality | Baseline | Follow-up |
|---|---|---|
| alimentary tract and metabolism | 144 | 145 |
| blood and blood forming organs | 12 | 22 |
| cardiovascular system | 124 | 142 |
| dermatologicals | 108 | 64 |
| genito urinary system and sex hormones | 72 | 76 |
| systemic hormonal preparations, excl. sex hormones and insulins | 26 | 24 |
| antiinfectives for systemic use | 56 | 35 |
| antineoplastic and immunomodulating agents | 5 | 5 |
| musculo-skeletal system | 145 | 183 |
| nervous system | 710 | 729 |
| antiparasitic products, insecticides and repellents | 5 | 4 |
| respiratory system | 721 | 538 |
| sensory organs | 42 | 42 |
| various | 1 | 3 |
